# VoxNeuro: Leakage-Safe Parkinson’s Screening Support via Multi-View Grassmannian Speech Analysis

**DOI:** 10.64898/2026.09.27.26364114

**Authors:** Laiba Khan, Manas Waghe

## Abstract

Parkinson’s disease (PD) affects speech production, motivating classification from repeated voice measurements. Record-wise evaluation can place recordings from the same participant in both training and test sets. We propose VoxNeuro, a novel subject-level framework combining Grassmann representations of repeated recordings with Euclidean mean-dispersion summaries through a provably positive-semidefinite kernel for support vector classification. Features undergo training-fold Gaussian rank normalization, and a supervised partial least squares subspace ensemble is activated when the feature count exceeds the number of training recordings. Two public cohorts were evaluated, UCI-489 (80 subjects) and PD-252 (252 subjects), each with three sustained-vowel recordings per subject. At the reference five-fold subject-disjoint partition, balanced accuracy was 87.5% and 81.6%, respectively, exceeding all five standard comparators under identical folds and normalization. PD-252 sensitivity was 89.9% and specificity was 73.4%. Across 20 additional subject partitions, PD-252 balanced accuracy averaged 78.4%, with a mean paired advantage of 3.2 percentage points over the highest-scoring standard comparator. VoxNeuro exceeded each standard comparator in at least 19 partitions for both balanced accuracy and macro-F1. Ablations favored rank normalization and the unaugmented supervised-subspace configuration on PD-252. VoxNeuro thus offers an effective and reproducible framework for subject-level analysis of repeated speech measurements in PD screening research.

## 1. Introduction

Parkinson’s disease (PD) is a progressive neurodegenerative disorder involving nigrostriatal dopaminergic loss, basal ganglia circuit dysfunction, and heterogeneous motor and non-motor manifestations [1–3]. Its diagnosis remains clinical [4,5], and its global burden has grown faster than that of any other neurological disorder [6,7]. Disordered speech is among the most common motor consequences: dysarthria, hypophonia, and related vocal-tract dysfunctions affect the large majority of patients over the disease course [8,9]. Quantitative acoustic analysis detects these changes even in early and untreated disease [10]. Nonlinear dysphonia measures separate disordered from healthy voices [11], and repeated noninvasive speech tests have supported research on low-burden remote monitoring and progression tracking [12–14]. Automated speech analysis has revealed prodromal changes in at-risk cohorts [15]. Machine-learning approaches to voice-based PD detection have accordingly expanded rapidly, as documented by recent systematic reviews [16,17]. A digital biomarker, however, is clinically credible only when its evaluation matches its intended unit of use: the patient.

Public PD speech datasets often contain multiple recordings per participant. Record-wise splitting can place recordings from the same participant in both training and test sets. Evaluation then combines predictions for participants represented in training with predictions for previously unseen participants, allowing subject-specific anatomical and acquisition signatures to influence the reported performance. This failure mode is a specific instance of data leakage, a recognized driver of over-optimistic results across applied machine learning [18,19]. Saeb et al. demonstrated that record-wise evaluation can yield higher accuracy than subject-wise evaluation of the same pipeline [20], prompting further discussion of cross-validation design [21]. Subject characteristics, model selection, and resampling design also affect performance estimation [22–27]. The UCI-489 cohort contains three recordings per subject [28], and replication-aware classification methods have been developed for this dataset [29].

Four methodological strands inform this study. First, engineered speech descriptors from the dysphonia, cepstral, wavelet, and tunable Q-factor wavelet transform (TQWT) families have been used to characterize PD-related voice changes [12,30,31]. Second, in subspace geometry, collections of related observations can be represented as points on the Grassmann manifold Gr(*r, d*), whose quotient structure removes arbitrary basis choices [32–34]. This representation underpins subspace-based learning in computer vision and pattern recognition [35,36], and kernels on such manifolds require care because Gaussian functions of arbitrary manifold distances are not automatically positive semidefinite (PSD) [37]. Third, in kernel fusion, reproducing-kernel Hilbert space (RKHS) theory [38–40] and multiple-kernel learning [41–43] combine heterogeneous similarities while preserving convex support vector optimization. Fourth, synthetic oversampling addresses class imbalance [44–46]; its behavior in high-dimensional feature spaces has been characterized in detail [47]. This study combines these strands under an evaluation protocol that treats the patient as the statistical unit.

Deep architectures, including convolutional, recurrent, and transformer-based models, are increasingly applied to voice-based PD detection [16]. Smartphone studies such as mPower illustrate opportunities for remote speech collection [48,49], consistent with technology roadmaps for PD assessment [50–52]. The present study examines two repeated-phonation cohorts containing 80 and 252 subjects, for which VoxNeuro combines subject-level subspace representations with kernel support vector classification.

In this work, we propose VoxNeuro, a novel subject-level framework that aligns the mathematical unit of analysis with the clinical unit of interest: each subject is represented by the complete set of repeated measurements. At the reference subject-disjoint partition, VoxNeuro achieves higher balanced accuracy than all five standard comparators on both public cohorts, exceeds each of them in at least 19 of 20 alternative partitions of the imbalanced cohort, and is reproducible from the archived code and predictions. A fixed-rank representation based on singular value decomposition (SVD) encodes the joint orientation of the repetitions, while a Euclidean summary preserves feature-space location and within-subject variability. Both views are built after fold-local rank normalization and, in the high-dimensional regime, inside a supervised low-dimensional subspace; a positive-semidefinite direct-sum kernel combines them, and evaluation is subject-disjoint throughout (Figure 1). For imbalanced full-space training folds, synthetic subjects are generated along manifold geodesics rather than by interpolating individual recording rows. A research-stage web prototype for guided speech capture complements the analytical work and is described in Section 4.

**Figure 1.**
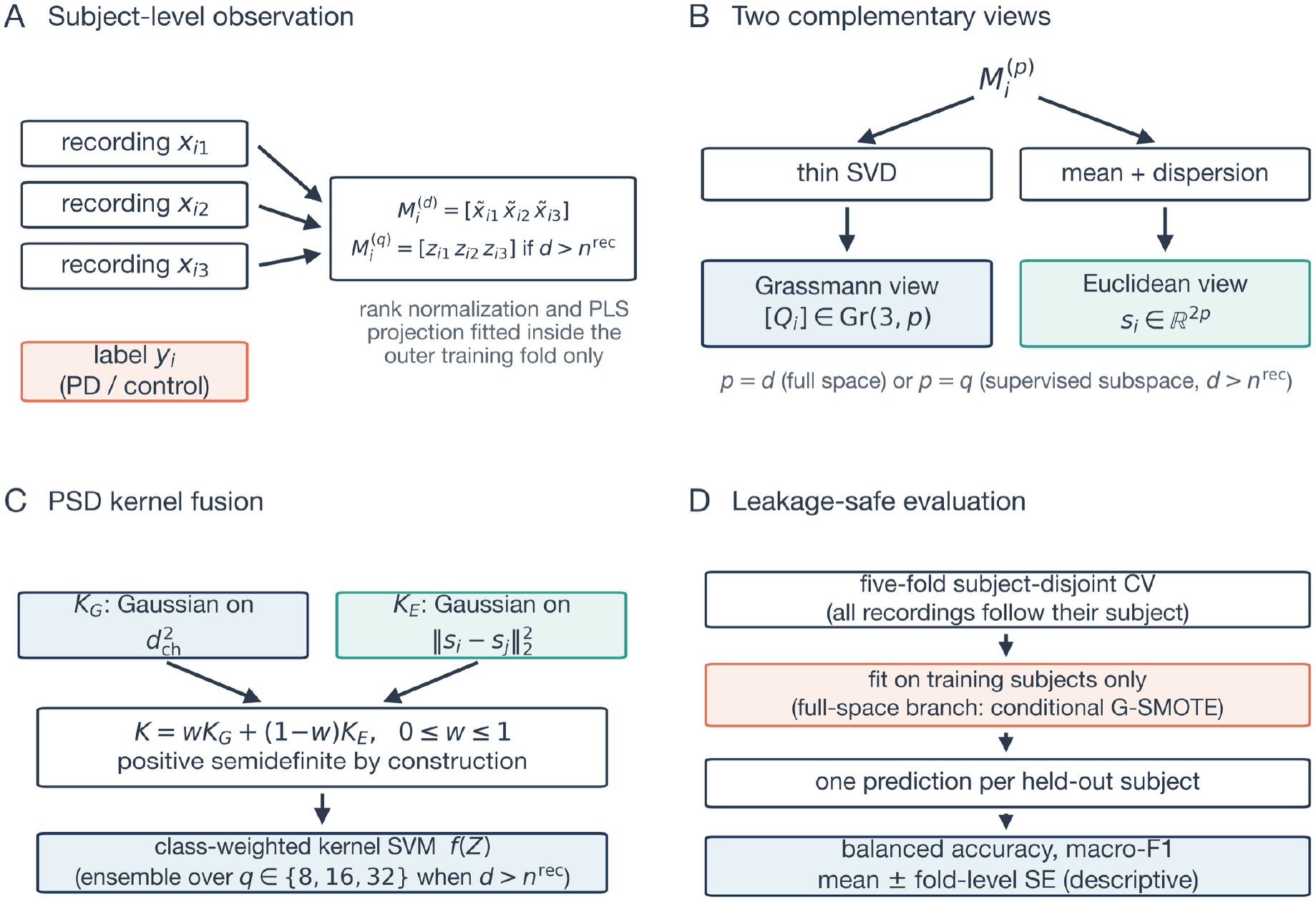
Schematic overview of VoxNeuro. (**A**) Each subject contributes three repeated recordings and one diagnostic label. The recordings are normalized with functions fitted inside the outer training fold only (Gaussian rank normalization) and stacked into a matrix *M*_*i*_; when the feature count exceeds the number of training recordings, the recordings are first projected onto a supervised subspace (Section 2.7). (**B**) A thin singular value decomposition (SVD) yields the Grassmann view [*Q*_*i*_] ∈ Gr(3, *p*), while coordinate-wise mean and dispersion yield the Euclidean view *s*_*i*_ ∈ ℝ^2*p*^, with *p* = *d* in the full-space branch and *p* = *q* in the supervised-subspace branch. (**C**) Gaussian kernels on the two views are combined into a positive-semidefinite (PSD) fused kernel feeding a class-weighted support vector machine (SVM). (**D**) Evaluation is five-fold subject-disjoint cross-validation (CV); in the full-space branch, conditional Grassmann-geodesic synthetic minority oversampling technique (G-SMOTE) is activated only inside imbalanced training folds, and fold scores are summarized as mean ± standard error (SE). PD, Parkinson’s disease.

The main contributions of this work are as follows. First, we define the subject-disjoint prediction target and express record-wise risk as a mixture of seen-person and unseen-person risks, identifying the condition under which subject overlap produces optimistic estimates. Second, we introduce a two-view kernel construction that fuses a basis-invariant Grassmann representation of repeated speech with a Euclidean mean–dispersion summary through a provably positive-semidefinite kernel. Third, we combine fold-local Gaussian rank normalization with an effective dimension-adaptive supervised-subspace ensemble that is activated only when the feature count exceeds the number of training recordings. Fourth, we implement conditional Grassmann-geodesic oversampling for imbalanced full-space training folds and characterize it in the full-space and oversampling ablations.

A shared configuration was evaluated against five standard comparators (class-weighted logistic regression, a class-weighted linear support vector machine (SVM), logistic regression after the synthetic minority oversampling technique (SMOTE), a class-weighted random forest, and partial least squares (PLS) discriminant analysis) and a full-space Euclidean-kernel ablation under identical folds and normalization. Ablations examined normalization, the subspace representation, view fusion, oversampling, and the gender covariate, while 20 alternative subject partitions assessed sensitivity to the evaluation split. The remainder of this paper is organized as follows. Section 2 describes the cohorts, the model, and the evaluation protocol; Section 3 reports the results; Section 4 discusses the findings, the capture prototype, and the limitations of the study; and Section 5 concludes.

## 2. Materials and Methods

### 2.1. Datasets

We analyzed two public, de-identified cohorts (Table 1). The first, denoted UCI-489, comprises 80 subjects (40 PD, 40 control) with three recordings of sustained /a/ per subject, each recording described by 44 replicated acoustic descriptors and one dataset-provided binary variable labeled gender [28,29]. The second, denoted PD-252 [53], comprises *2*5*2* subjects (188 PD, 64 control) with three recordings per subject, described by 75*2* descriptors spanning baseline acoustic, vocal/time-frequency, Mel-frequency cepstral coefficient (MFCC) [54], wavelet, TQWT [55], and intrinsic mode function/empirical mode decomposition (IMF/EMD) [56] families [30,31]. It also includes one dataset-provided binary variable labeled gender, which constitutes the demographic family of the permutation analysis in Appendix A.*2*. We retain the source label for this binary input variable; it was included as a predictor, and fold construction used diagnostic labels and subject identifiers only. It is unequally distributed across the diagnostic classes (code 1 in 14 of 40 PD and 18 of 40 control subjects in UCI-489, and in 107 of 188 PD and *2*3 of 64 control subjects in PD-*2*5*2*), and its contribution to performance is quantified in Section 3.3. UCI-489 provides a balanced, lower-dimensional setting in which VoxNeuro uses the full-space representation. PD-*2*5*2* provides an imbalanced, higher-dimensional setting in which VoxNeuro uses the supervised-subspace ensemble (Section 2.7); the full-space ablations use conditional G-SMOTE.

**Table 1.** Dataset regimes and evaluation design.

| Attribute | UCI-489 | PD-252 |
| --- | --- | --- |
| Subjects | 80 | 252 |
| PD / control | 40 / 40 | 188 / 64 |
| Recordings per subject | 3 | 3 |
| Modeled variables | 45 | 753 |
| Recording task | Sustained /a/, three repetitions | Sustained /a/, three repetitions |
| Descriptors | 44 replicated acoustic descriptors + gender | 752 multi-family engineered descriptors + gender |
| Imbalance handling | Class weighting (balanced folds) | Class weighting for VoxNeuro, logistic regression, linear SVM, and random forest; equal class priors for PLS-DA; ordinary SMOTE for SMOTE + logistic regression; G-SMOTE for full-space kernel models |
| Outer evaluation | Five-fold subject-disjoint cross-validation |  |

G-SMOTE, Grassmann-geodesic synthetic minority oversampling (Section 2.8); SMOTE, synthetic minority oversampling technique; PLS-DA, partial least squares discriminant analysis.

**Table 2.**
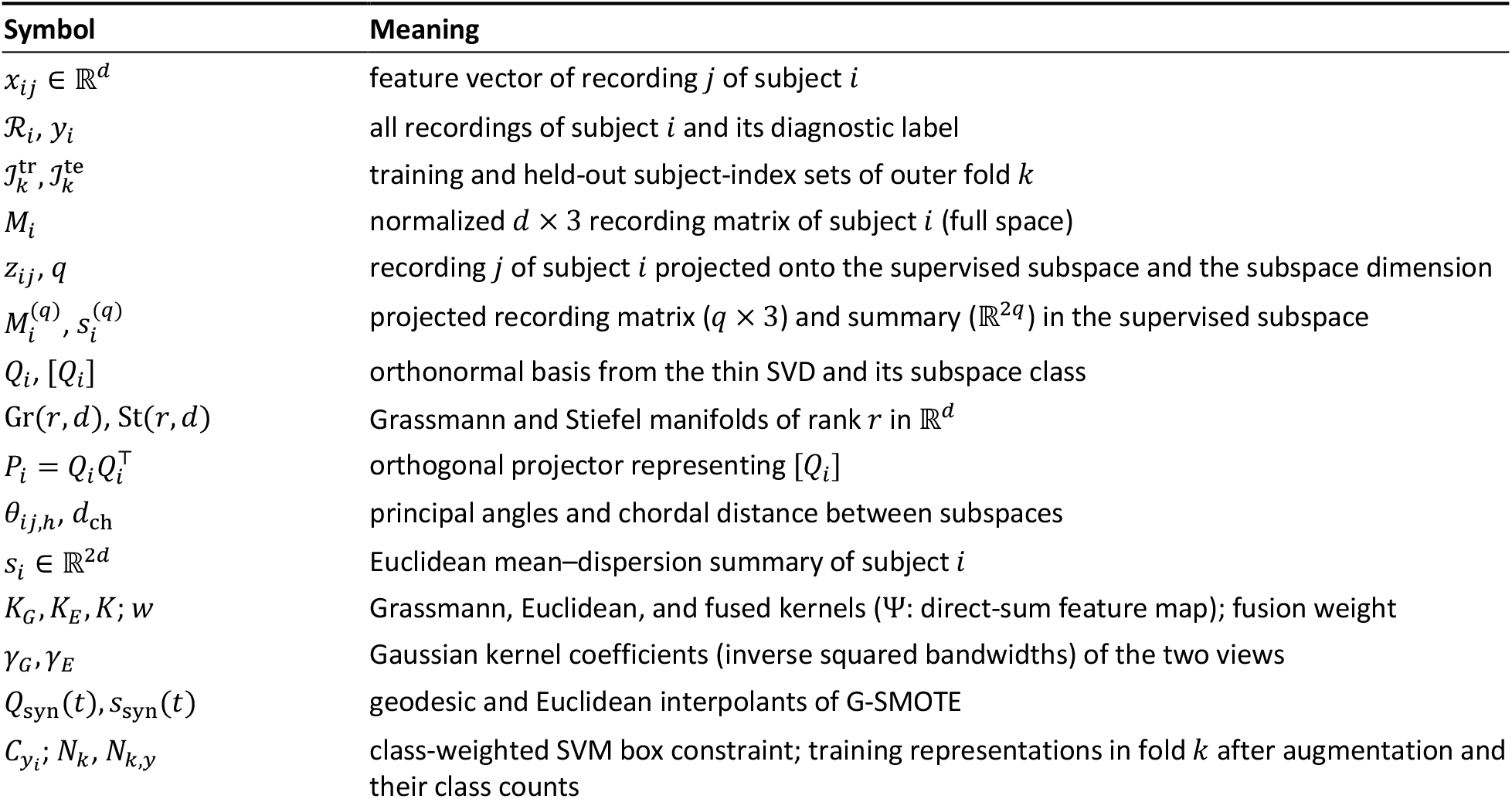
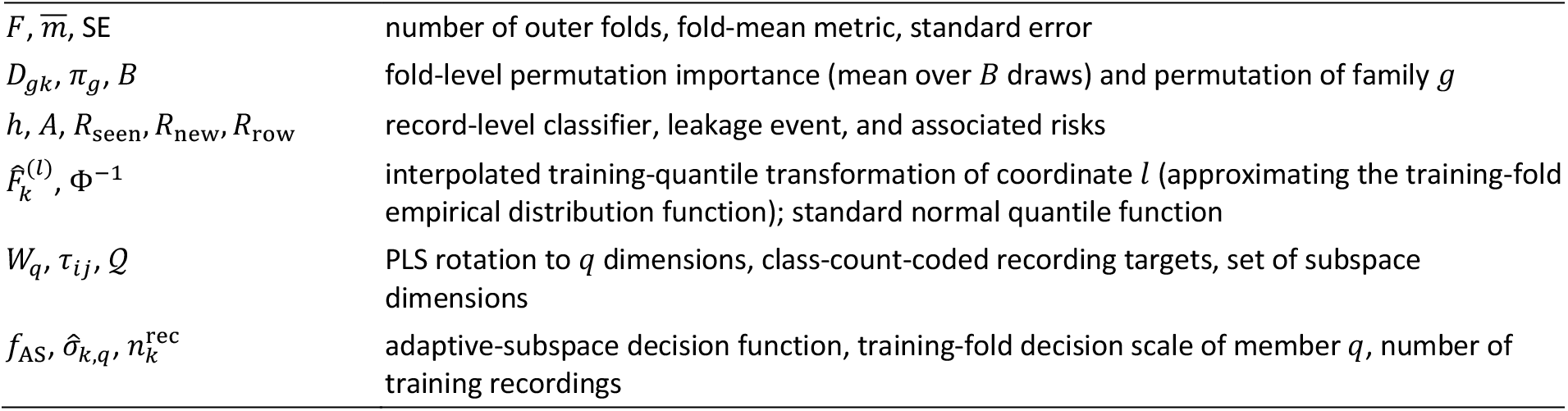
Principal notation.

### 2.2. Subject-Disjoint Problem Formulation

Let ℛ_*i*_ = {(*x*_*ij*_, *y*_*i*_): *j* = 1, …, *m*_*i*_} denote all recordings from subject *i*; Table 2 summarizes the notation used throughout. For outer fold *k*, let 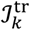 and 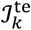 be the training and held-out subject-index sets. Leakage-safe evaluation requires

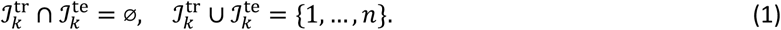

All recordings in ℛ_*i*_ are assigned to the same partition as subject *i* (Figure 2A,B). For a pointwise classification loss *l* and the model *f*_*k*_ fitted on the training subjects of fold *k*, the fold risk is evaluated once per held-out subject,

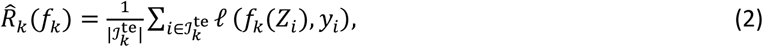

where *Z*_*i*_ is the complete subject-level representation. Equations (1) and (*2*) define the intended estimand: generalization to an unseen person rather than to an unseen row from a known person. The reported balanced accuracy and macro-F1 are fold-level aggregate metrics defined in Equation (17).

**Figure 2.**
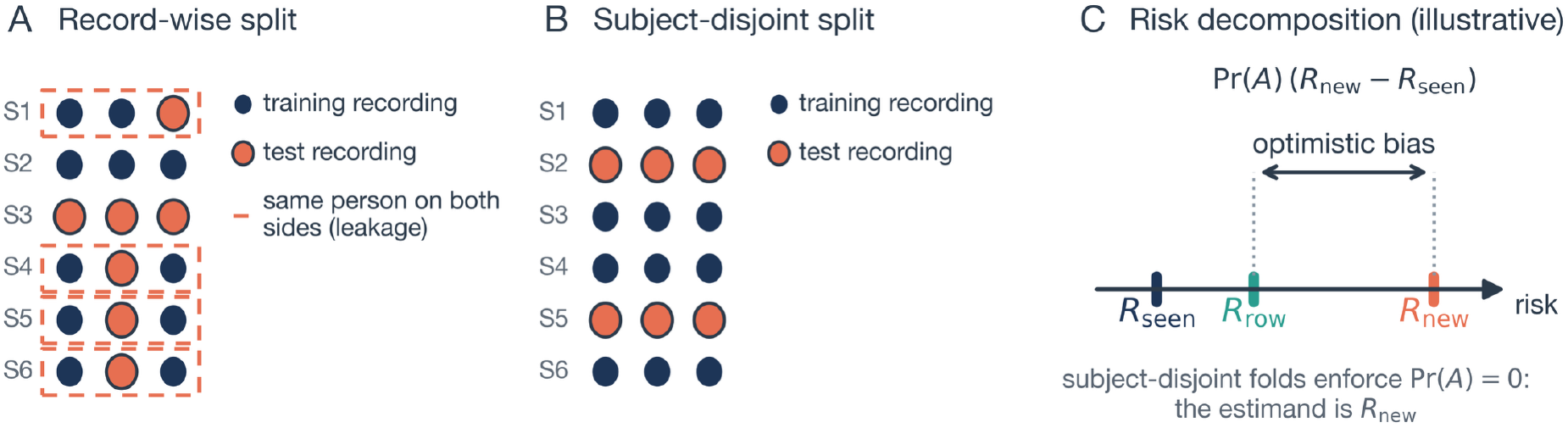
Schematic illustration of evaluation leakage with repeated recordings. (**A**) A record-wise split assigns individual recordings to training or test sets, so several subjects (dashed outlines) appear on both sides and the test set is not independent of training identities. (**B**) A subject-disjoint split assigns all recordings of a subject jointly. (**C**) Illustration of Equation (20) for *R*_*seen*_ < *R*_*new*_ and Pr(*A*) = 0.65: the record-wise risk *R*_*row*_ is a mixture of the seen-person risk *R*_*seen*_ and the unseen-person risk *R*_*new*_, and the arrow marks the optimistic bias Pr(*A*) (*R*_*new*_ − *R*_*seen*_).

### 2.3. Normalization and Subject Representation

For subject *i*, the three recording-level feature vectors *x*_*ij*_ ∈ ℝ^*d*^ are normalized with functions estimated exclusively from the recordings of training subjects in outer fold *k*. VoxNeuro uses Gaussian rank normalization. For coordinate *l*, let 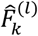 denote the interpolated training-quantile transformation of the training-fold recordings, and let Φ^−1^ denote the standard normal quantile function, with its argument clipped to [10^−7^, 1 − 10^−7^]:

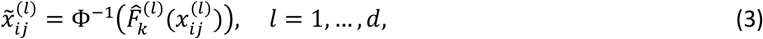

The transformation 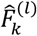 approximates the training-fold empirical distribution function through 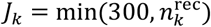quantile knots with linear interpolation between them, where 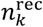 is the number of training recordings (*J*_*k*_ = 192 on UCI-489 and 300 on PD-252); tied values share one image. It is monotone in every coordinate and is fitted on training recordings only; hel2d-out recordings are mapped through the frozen training-fold functions. Many engineered acoustic descriptors are heavy-tailed; the rank transformation is fitted without diagnostic labels and expresses each coordinate on a Gaussian scale that is robust to heavy-tailed marginals before subject representation. The binary gender covariate maps to two values near ±5.2 (compared with about ±1 under z-standardization), and in the supervised-subspace branch it enters the fitted PLS projection with the other recording-level features; its contribution is quantified in Section 3.3. Z-standardization with a training-fold mean and scale vector (zero-variance coordinates assigned unit scale) is retained as an ablation (Section 2.11). In both cohorts every subject has *m*_*i*_ = 3 recordings, so the normalized recordings are stacked column-wise into a *d* × 3 matrix, and a thin SVD [57] gives

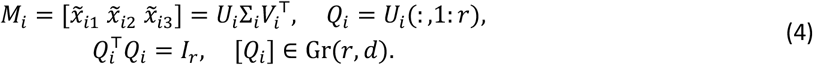

Throughout, the rank is fixed at *r* = 3, matching the three repeated recordings per subject. The basis is not unique: *Q*_*i*_ and *Q*_*i*_*O*, with *O* in the orthogonal group O(*r*), represent the same subspace. This quotient structure removes coordinate choices within the retained SVD basis (Figure 3A). In the public releases of both datasets, one subject per cohort (CONT-36 in UCI-489; subject 37 in PD-252) contains byte-identical repeated recordings, so its matrix has numerical rank two. For these subjects, the implementation retains the third orthonormal vector supplied by the SVD under an explicit rank-completion option. In the high-dimensional regime defined in Section 2.7, the same construction is applied to supervised low-dimensional projections of the recordings.

**Figure 3.**
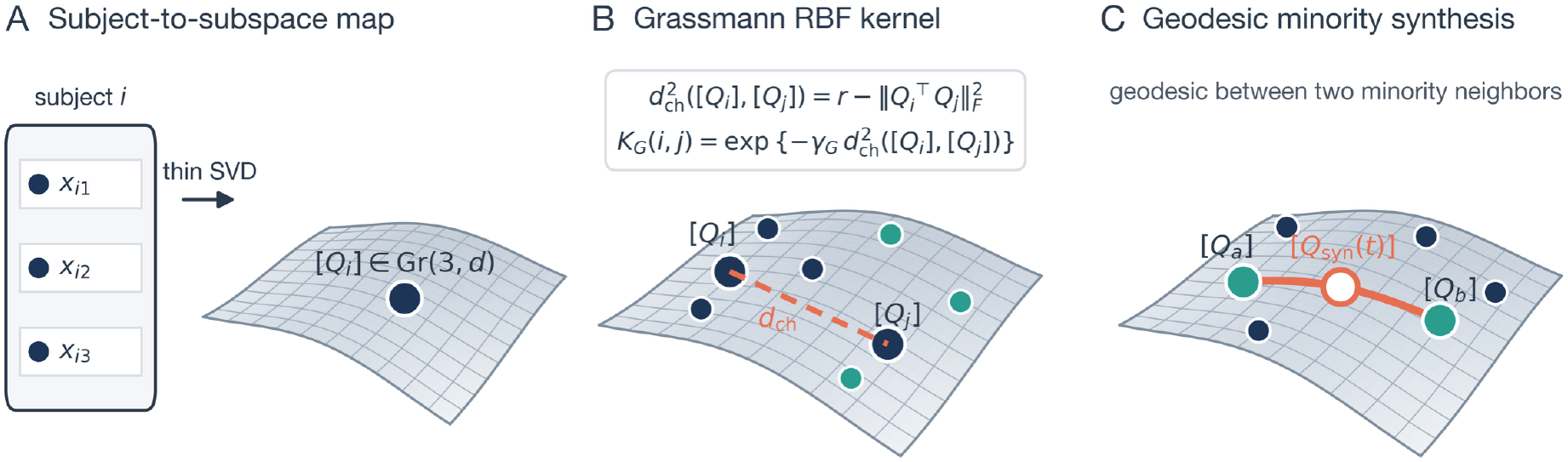
Geometric interpretation of the VoxNeuro representation (schematic; the curved sheet stands for the Grassmann manifold). (**A**) In the full-space construction, the three repeated recordings of a subject are mapped by a thin SVD to a point [*Q*_*i*_] on Gr(3,*d*); the supervised-subspace construction uses Gr(3,*q*). (**B**) The chordal distance *d*_*ch*_ between two subject subspaces (dashed chord), whose square enters the displayed Gaussian Grassmann radial-basis-function kernel, compares subjects independently of the arbitrary orthonormal basis selected for each subspace; navy and teal points denote subjects of the two classes. (**C**) Schematic of minority synthesis along the Grassmann geodesic joining two minority neighbors, rather than row-wise Euclidean interpolation.

### 2.4. Principal Angles and Projection Geometry

Principal angles between two subject subspaces (Figure 4A) are defined by [32,58]

**Figure 4.**
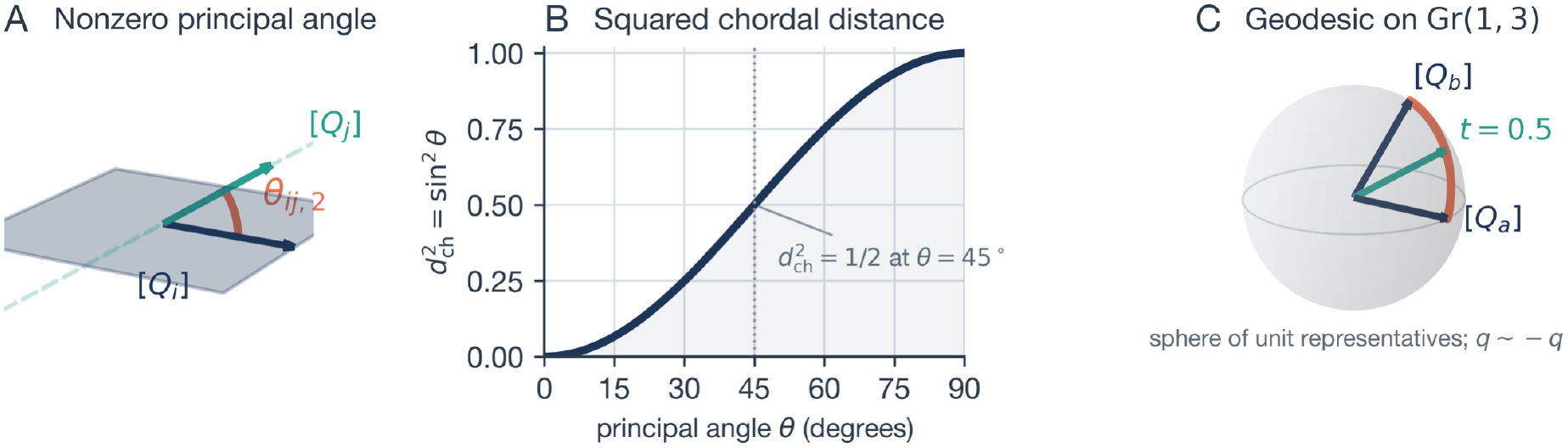
The geometry behind Equations (5), (6), (13), and (14), drawn for illustrative toy subspaces. (**A**) Two planes in ℝ^3^ that share one direction, so the first principal angle is zero and the drawn angle is the second principal angle *θ*_*ij*,2_. (**B**) Exact rank-one relation between a principal angle and the squared chordal distance, 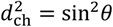. (**C**) A geodesic path computed with Equations (13) and (14) between two one-dimensional subspaces [*Q*_*a*_] and [*Q*_*b*_]; every interpolated frame in the drawn path satisfies *Q*_syn_(*t*)^T^*Q*_syn_(*t*) = *I*_*r*_ to machine precision, consistent with Proposition 2.

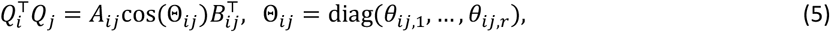

where *A*_*ij*_ and *B*_*ij*_ are orthogonal. With the projector 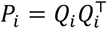, the squared chordal distance is given by [33,59]

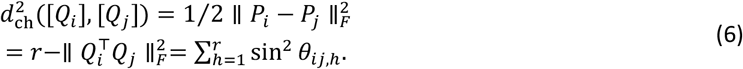

Because (*Q*_*i*_*O*)(*Q*_*i*_*O*)^T^ = *P*_*i*_, the distance in Equation (6) is invariant to the choice of orthonormal basis. Vectorizing *P*_*i*_ maps the chordal distance to a Euclidean distance in projector space, up to a constant factor 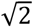 absorbed by the kernel coefficient, thereby providing a Euclidean embedding for the Gaussian Grassmann kernel.

### 2.5. Complementary Euclidean View

The complementary Euclidean view retains the coordinate-wise mean and dispersion:

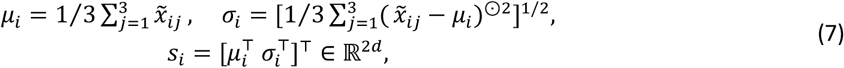

where (⋅)^⊙2^ and the square root are applied elementwise. Thus [*Q*_*i*_] encodes the orientation of the span of the normalized repetitions (*M*_*i*_ is not centered within subject, so this span generally includes the mean direction), whereas *s*_*i*_ retains coordinate-level location and dispersion.

### 2.6. Direct-Sum Kernel and Guarantee of Positive Semidefiniteness

Let *ϕ*_*G*_ and *ϕ*_*E*_ be feature maps for Gaussian kernels on the projection and Euclidean views [38,39]. With fusion weight *w*, the joint representation is the Hilbert direct sum

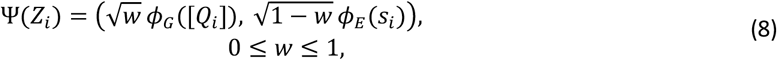

which induces, for kernel coefficients *γ*_*G*_, *γ*_*E*_ > 0 (inverse squared bandwidths),

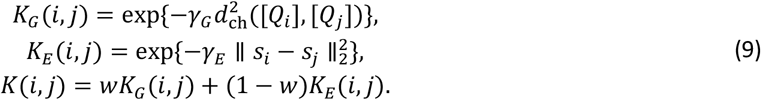

#### Proposition 1

*(basis invariance and positive semidefiniteness)*.

The kernel in Equation (9) is invariant to *Q*_*i*_ ↦ *Q*_*i*_*O* and is PSD. Basis invariance follows from the projector representation in Equation (6). Each Gaussian block is PSD on its Euclidean embedding, and a nonnegative weighted sum of PSD kernels is PSD [39,41]; hence, for every *c* ∈ ℝ^*n*^,

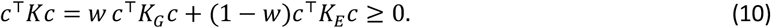

Equivalently, *K*(*i, j*) = ⟨Ψ(*Z*_*i*_), Ψ(*Z*_*j*_)⟩ in the direct-sum reproducing-kernel Hilbert space. □ The projector embedding is essential. Gaussian kernels built on other manifold distances, such as the geodesic arc length, are not PSD for all bandwidths [37], whereas the chordal construction used here inherits positive semidefiniteness from its explicit Euclidean embedding. Basis invariance applies to alternative orthonormal bases of a fixed retained subspace.

### 2.7. Dimension-Adaptive Supervised Subspace

VoxNeuro constructs supervised low-dimensional subject representations when the feature count exceeds the number of training recordings, a regime in which high-dimensional subspace distances can concentrate and reduce the variation of kernel similarities; the projection is designed to restore distance dispersion and improve kernel discrimination in this regime (Section 3.3). The projection is a PLS regression [60,61] of the normalized recordings on class-count-coded binary targets 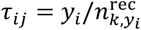 . Here *y*_*i*_ = +1 denotes PD and *y*_*i*_ = −1 denotes control, each recording carries its subject’s label, and 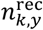 is the number of training recordings with label *y*. For this single-response regression the coding is an affine recoding of the binary labels; class imbalance is handled by the SVM class weights (Section 2.9). The regression was fitted without target or predictor scaling; with *w*_*q*_ ∈ ℝ^*d*×*q*^ the fitted rotation matrix and 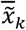 the mean of the normalized training recordings, every recording is projected as

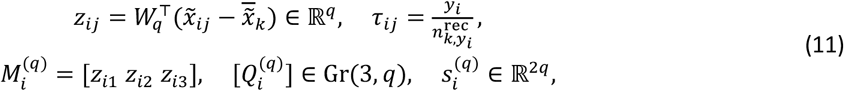

and the subject views of Sections 2.3–2.5, the median bandwidth rule, and the fused kernel of Equation (9) are then built in ℝ^*q*^ exactly as in the full space, with the shared weight *w* = 0.5. The projection is fitted separately within each outer training fold; distance-dispersion diagnostics for the full-space and projected representations are reported in Section 3.3. One class-weighted SVM *f*_*q*_ is trained for each dimension *q* ∈ *Q* = {8,16,32}. Instead of selecting a dimension within the fold, the decision values of the three members are divided by their training-fold standard deviations 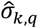 and averaged, giving a fixed-dimension ensemble; 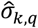 is the population standard deviation of the member’s decision values over the training subjects, whose mean is 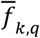, plus 10^−12^:

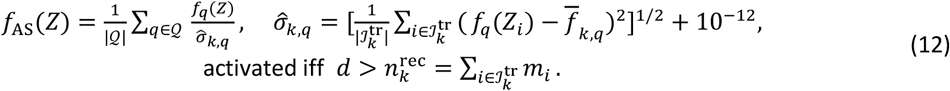

The stage is activated by a fixed dimensionality gate: it is applied only when *d* exceeds the number of training recordings 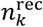, which holds on PD-252(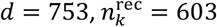or 606) and not on UCI-489 (*d* = 45, 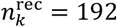). The gate, the dimension set, and the equal-weight average are part of the shared configuration of Section 2.11. When the gate is inactive the model reduces to the full-space fused model of Equation (9) under rank normalization, including the conditional geodesic oversampling branch (G-SMOTE, Section 2.8) for imbalanced training folds. Inside the supervised subspace no synthetic subjects are generated; the class-weighted SVM of Section 2.9 accounts for class imbalance through class-specific penalties, and the corresponding oversampling ablation is reported in Section 3.3. The complete model is referred to as VoxNeuro. Its ablations are the rank-normalized full-space model (no subspace stage), the z-standardized full-space model, the full-space Euclidean-kernel ablation (*w* = 0), and the view ablations (*w* = 0 and *w* = 1 with the automatic gate retained).

### 2.8. Grassmann-Geodesic Synthetic Minority Oversampling

In the full-space branch, imbalanced training folds are balanced by a project-specific manifold adaptation of synthetic oversampling, referred to as Grassmann-geodesic SMOTE (G-SMOTE); the abbreviation does not denote the separate Geometric SMOTE algorithm of Euclidean imbalanced learning [62]. Following the synthetic-neighbor principle of standard SMOTE [44] and its descendants [45,46,63], for minority subjects *a* and *b*,

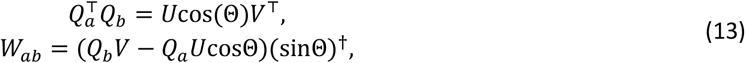

where † denotes the Moore–Penrose pseudoinverse. A shortest geodesic and its coupled Euclidean interpolation are defined as follows (Figure 4C):

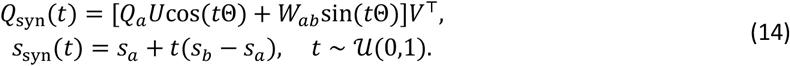

Within each imbalanced training fold, the minority class was the control class. For each synthetic subject, one minority subject was sampled uniformly at random, and a partner was selected uniformly from that subject’s five nearest minority neighbors under squared chordal distance. Synthetic subjects were generated until the two class counts were equal. The same sampled value of *t* was used for the Grassmann and Euclidean interpolations. For numerical stabilization, singular values were clipped to [−1,1] before the arccosine, directions with sin*θ* ≤ 10^−10^ were treated as zero-angle directions, and the implementation applied a QR factorization to *Q*_syn_(*t*). The resulting invertible right basis change preserves the represented Grassmann point. No synthetic subjects were generated for balanced UCI-489. Interpolating whole-subject representations along the manifold avoids creating synthetic recording rows and selects partners by subspace geometry.

#### Proposition 2

*(manifold validity)*

In exact arithmetic, the nonzero-angle columns of *Q*_*a*_*U* and *w*_*ab*_ are mutually orthonormal, while zero-angle directions remain in the cosine term. Consequently,

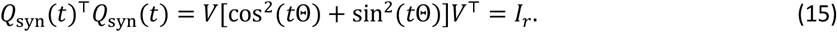

Therefore, *Q*_syn_(*t*) ∈ St(*r, d*), the Stiefel manifold of orthonormal *d* × *r* frames; it represents a valid Grassmann point. Its endpoints are equivalent to [*Q*_*a*_] and [*Q*_*b*_]. □ Proposition *2* establishes the validity of the Grassmann component; Equation (14) defines the accompanying Euclidean summary by linear interpolation.

### 2.9 Kernel Support Vector Machine, Metrics, and Complexity

For SVM training [64], let 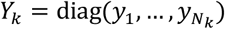 over the training representations of fold *k*. Balanced class weighting sets 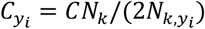, where *N*_*k*_ is the number of training representations in outer fold *k* after any training-only augmentation and *N*_*k,y*_ is the corresponding class count; this is the balanced heuristic implemented in standard open-source libraries [65]. When the training classes are exactly balanced, as on UCI-489 and after G-SMOTE parity on PD-252, these weights reduce to *C*_*yi*_= *C*. Class weighting is therefore active for the supervised-subspace SVMs of VoxNeuro and for the class-weighted logistic regression, linear SVM, and random forest comparators on PD-252; PLS-DA uses equal class priors (Section 2.11). The class-weighted dual and the decision function with bias *b* are

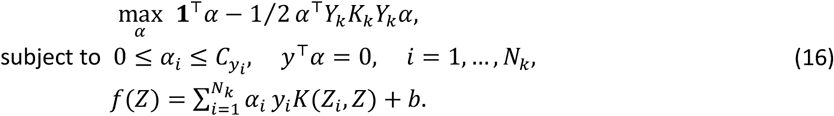

Balanced accuracy (BA) [66] and macro-F1 [67] weight diagnostic classes symmetrically:

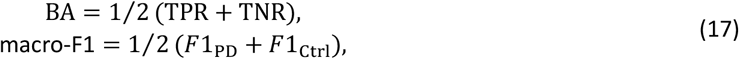

where TPR is sensitivity, TNR is specificity, and *F*1_PD_ and *F*1_Ctrl_ are the class-wise F1 scores. Balanced accuracy averages the two class recalls, whereas macro-F1 averages the two class-specific F1 scores; the Matthews correlation coefficient is a common alternative [68]. For *F* = 5 outer folds, a metric *m* is summarized by

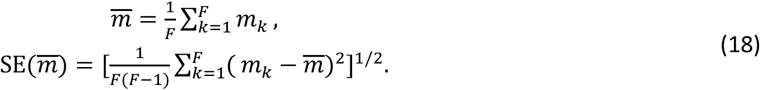

The standard error (SE) across folds summarizes the fold-to-fold variability of the cross-validation estimate. Balanced accuracy and macro-F1 were summarized across folds with Equation (18), whereas sensitivity, specificity, and the predictive values were computed from pooled out-of-fold confusion counts. All of these metrics are reported in percent, and differences between models are reported in percentage points.

For *n* subjects and rank *r* = 3, subject SVDs require *O*(*ndr*^2^), the Grassmann Gram matrix requires *O*(*n*^2^*dr*^2^), and the Euclidean block requires *O*(*n*^2^*d*). These bounds cover view construction only and exclude the kernel-SVM optimization, which can dominate as *n* grows. The subspace branch additionally requires three PLS fits and projections, three kernel constructions, and three kernel-SVM optimizations, and the full-space branch adds the G-SMOTE neighbor search where active. All components remain tractable for the present cohort sizes *n* = 80 and *n* = 252.

### 2.10. Formal Effect of Record-Wise Leakage

The mechanism illustrated in Figure 2 can be stated formally, following record-wise versus subject-wise analyses in clinical machine learning [18,20,21]. Let (*X, y*) be a random validation recording and its label, let *A* be the event that the recording belongs to a person represented in training, and let *h* be a record-level classifier. With *A*^*c*^ the complement of *A*, define

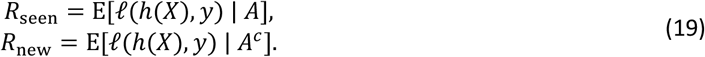

A record-wise split then estimates

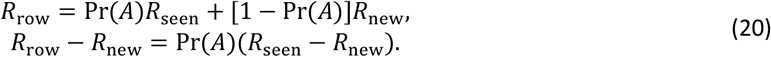

If repeated recordings from previously observed people are easier to classify than recordings from unseen people, then *R*_seen_< *R*_new_, and the record-wise estimate is optimistically biased in direct proportion to Pr(*A*) (Figure 2C). Subject-disjoint folds enforce Pr(*A*) = 0 by construction.

### 2.11. Experimental Protocol, Comparators, and Ablations

Outer folds were generated by five-fold stratified subject-level cross-validation with shuffling and a random seed of 42, referred to as the reference partition (Figure 5A). For each outer fold, the recording-level normalizer was fitted only to recordings from training subjects. Each kernel coefficient was estimated from the positive squared pairwise distances δ^2^ of the corresponding view over the training subjects (after G-SMOTE augmentation where active) as *γ* = [2 median(δ^2^) + 10^−12^]^−1^. A single final configuration, *r* = 3, *w* = 0.5, *C* = 1, and *Q* = {8,16,32}, was applied to both cohorts. This configuration was fixed after development on the reference partition of both cohorts and was then held constant for the partition-sensitivity analysis described below. Within each training fold of the full-space branch, resampling was governed solely by the observed class counts. If the counts were unequal, the minority class was raised to the majority count using G-SMOTE for the full-space kernel models and ordinary SMOTE only for the comparator explicitly identified as SMOTE-based; if the counts were equal, resampling was the identity operation. All UCI-489 training folds were balanced, so no synthetic subjects were generated there. On PD-252 the full-space kernel models activated the branch in every fold, whereas VoxNeuro, whose gate activates the supervised-subspace branch on PD-252, generated no synthetic subjects on either cohort and is therefore deterministic given the outer partition. Table 3 summarizes the shared configuration, and the implementation is available in the project repository (see the Data Availability Statement).

**Table 3.** Final shared analysis configuration. For each fixed configuration, normalization functions, PLS projections, kernel coefficients, and classifier parameters are fitted using the outer training data.

| Component | Setting |
| --- | --- |
| Outer evaluation | five-fold stratified subject-level cross-validation, shuffled, seed 42 |
| Normalization | Gaussian rank normalization fitted on training recordings ($\min(300, n_k^{\text{rec}})$ quantile knots); z-standardization (training-fold mean/scale, unit scale for zero-variance coordinates) as an ablation |
| Supervised subspace | PLS projection fitted on training recordings with class-count-coded binary targets; members $q \in \{8, 16, 32\}$ averaged after division by the training-fold standard deviation of the decision values; activated only when $d$ exceeds the number of training recordings (PD-252: $753 > 603$ or $606$; UCI-489: $45 < 192$); no synthetic subjects in this branch |
| Subject representation | rank $r = 3$ thin SVD (Grassmann view) + mean/dispersion vector (Euclidean view) |
| Kernel coefficients | $\gamma = [2 \text{ median}(\delta^2) + 10^{-12}]^{-1}$ from positive squared pairwise training distances (augmented training set in the full-space branch on PD-252) |
| Fusion weight / penalty | $w = 0.5$; class-weighted SVM with $C = 1$; one shared post-development setting for both cohorts |
| Resampling rule | training-fold class-count gate in the full-space branch: identity when balanced (all UCI-489 folds); G-SMOTE to class parity when imbalanced, five nearest minority neighbors under chordal distance; the supervised-subspace branch uses class weighting only |
| Comparators | standard comparators: PLS-DA (components by inner cross-validation), class-weighted random forest (500 trees), class-weighted logistic regression, class-weighted linear SVM, SMOTE + logistic regression where resampling is active; full-space Euclidean-kernel ablation (fusion weight 0); all under rank normalization on identical folds |
| Metrics | balanced accuracy and macro-F1, mean ± fold-level SE; pooled out-of-fold confusion counts |
| Ablations and sensitivity | z-standardization; no subspace stage; $w = 0$ and $w = 1$ view ablations with the automatic gate (subspace ensemble on PD-252, rank-normalized full-space model on UCI-489); single subspace dimensions 4–64; gate forced on (UCI-489); G-SMOTE inside the subspace branch (24 seeds); removal of the gender covariate (reference partition and 20 partitions); 20 outer subject partitions for every model; full-space branch: 24 augmentation seeds, rank $r = 2$, augmentation family, fusion-weight sweep, feature-family permutation (Appendix A) |

**Figure 5.**
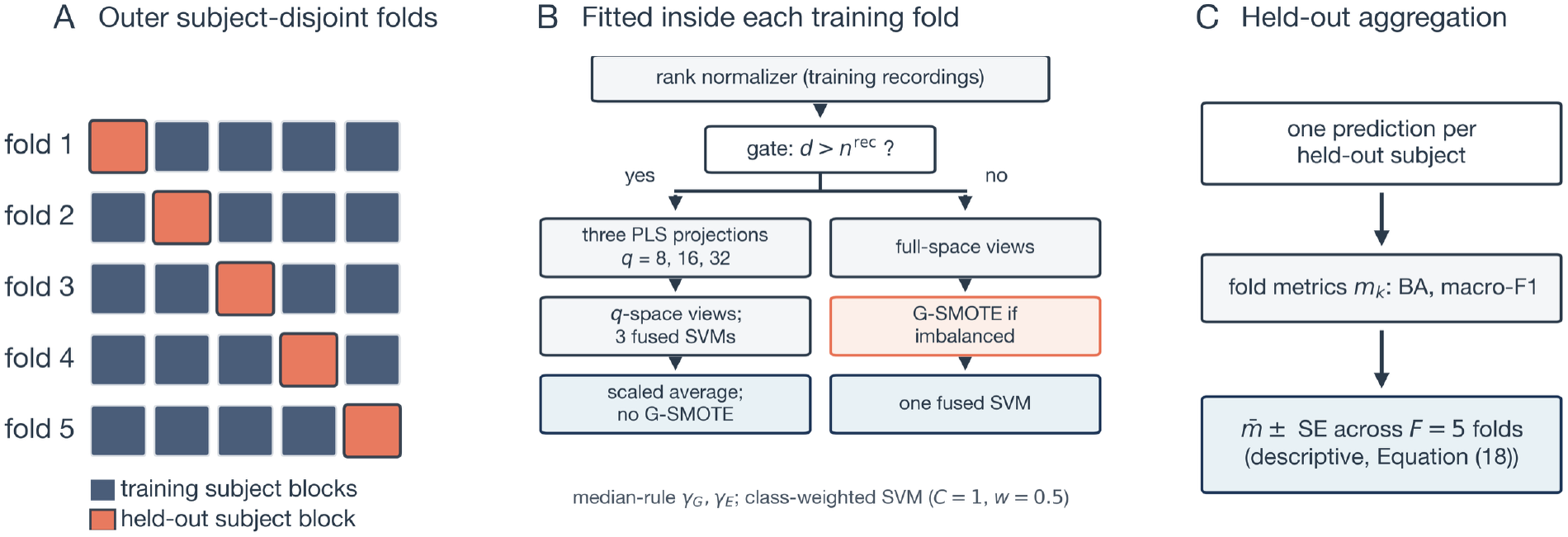
Evaluation protocol. (**A**) Five-fold subject-disjoint partitioning holds out one subject block (orange) in each evaluation fold. (**B**) Normalization functions, PLS rotations, kernel coefficients, and SVM parameters are fitted using the training subjects. After the rank normalizer, either the supervised-subspace branch (dimensionality gate active: PLS projections, *q*-space views, three SVMs averaged after scaling; no synthetic subjects) or the full-space branch (gate inactive: full-space views, conditional G-SMOTE in imbalanced folds, one SVM) is applied, with median-rule kernel coefficients in both branches. (**C**) Exactly one prediction is issued per held-out subject, fold metrics are computed at the subject level, and Equation (18) aggregates them across folds.

We compared VoxNeuro with five standard comparators trained on the same subject-level summary *s*_*i*_ under the same training-fold rank normalization and identical folds. They were class-weighted logistic regression, a class-weighted linear SVM, SMOTE followed by logistic regression where resampling is active, a class-weighted random forest (500 trees, ordinary bootstrap sampling, square-root feature subsampling, unrestricted depth, class weights recomputed within each bootstrap sample), and partial least squares discriminant analysis (PLS-DA). Logistic regression used L2 regularization with *C* = 1 (liblinear solver, at most 5000 iterations), and the linear SVM used the L2 penalty with squared-hinge loss and *C* = 1 (at most 20,000 iterations). SMOTE used five neighbors, or one fewer than the minority count when necessary. A subject was labeled PD when its decision score was strictly positive, and a forest probability of exactly 0.5 was assigned to control. For PLS-DA, the subject summaries were standardized with the training subjects, projected by PLS regression on the same class-count-coded targets without scaling to *q* components, and classified by shrinkage linear discriminant analysis with equal priors. The dimension *q* ∈ {2,4,8,16} was chosen by pooled inner five-fold out-of-fold balanced accuracy inside each training fold, with the smallest *q* retained on ties, and the scaler, projection, and discriminant were then refitted on the complete training fold. Inner selection was conditional on the recording-level normalizer fitted on the complete outer-training fold: the summary scaler, projection, and discriminant were refitted within each inner split, whereas the recording-level normalizer was not. A full-space Euclidean-kernel ablation with fusion weight *w* = 0 shares the summary features, the resampling, and the kernel family of the full-space model and differs from it only through setting the Grassmann kernel weight to zero in Equation (9). On PD-252 it retains the G-SMOTE branch and is therefore reported as an ablation rather than as a standard comparator. All comparators were implemented in the released codebase and verified against their stated configurations. We evaluated four groups of ablations. The first group concerns normalization and compares the rank-normalized and z-standardized full-space models. The second concerns the subspace stage: it is removed (rank-normalized full-space model), applied with the gate forced on for UCI-489 where the rule deactivates it, compared member by member against the three-member ensemble using *q* ∈ {4,8,16,32,64} on PD-252 and *q* ∈ {4,8,16,32} on UCI-489, and combined with geodesic oversampling inside the subspace branch across 24 augmentation seeds, where each ensemble member’s decision scale was computed over its complete augmented training set. The third concerns the views: *w* = 0 (Euclidean only) and *w* = 1 (Grassmann only) with the automatic dimensionality gate retained, so that these view ablations evaluate the supervised-subspace ensemble on PD-252 and the rank-normalized full-space model on UCI-489. The fourth removes the dataset-provided gender covariate from the feature set of VoxNeuro, the five standard comparators, and the rank-normalized full-space kernel models, at the reference partition and across the 20 alternative partitions. Supplementary analyses of a z-standardized full-space variant (augmentation-seed sensitivity, rank *r* = 2, augmentation family, fusion-weight sweep, and feature-family permutation sensitivity) are reported in Appendix A.

To assess the stability of the results, we repeated the entire five-fold evaluation for 20 alternative stratified subject partitions (StratifiedKFold seeds 0–19) with every model and summarized paired differences across partitions. Paired differences are reported in percentage points with one decimal, or with two decimals when their magnitude is below one percentage point. For models with the G-SMOTE branch the augmentation seed is derived from the partition seed, and for the other stochastic comparators the estimator or resampling random state also changes with the partition seed, so their summaries combine partition sensitivity with algorithmic randomness. VoxNeuro itself is deterministic conditional on the partition, and the archived fixed-augmentation partition results of the z-standardized full-space variant serve as an additional check. All analyses used Python 3.11.15 with NumPy 1.26.4, SciPy 1.11.4, pandas 2.0.3, scikit-learn 1.3.2, imbalanced-learn 0.11.0, and threadpoolctl 3.6.0 under single-threaded linear algebra. The released implementation and the archived subject-level predictions support regeneration of the reported analyses under this pinned environment, as detailed in the Data Availability Statement.

## 3. Results

### 3.1. Performance at the Reference Partition

VoxNeuro achieved the highest balanced accuracy and macro-F1 among the models in Table 4 at the reference partition (Figure 6). On UCI-489, balanced accuracy was 87.5% (SE 4.0%) and macro-F1 was 87.3% (SE 4.0%), compared with 82.5% and 82.2% for PLS-DA, the highest-scoring standard comparator. On PD-252, VoxNeuro achieved 81.6% (SE 4.0%) and 81.1% (SE 3.4%), compared with 77.7% and 78.6% for SMOTE plus logistic regression. The full-space Euclidean-kernel ablation reached 86.3% and 86.0% on UCI-489 and 76.6% and 75.7% on PD-252. Section 3.3 reports the normalization, subspace, and view ablations.

**Table 4.** Subject-level performance at the reference partition (mean ± fold-level standard error over five subject-disjoint folds, in percent). All models except the z-standardized full-space model share the same training-fold rank normalizer and outer folds; kernel models use the median-bandwidth rule and *C* = 1, whereas the other comparators retain their model-specific fitting procedures. Rows list the full model, its ablations, and the standard comparators. On UCI-489 the subspace stage is inactive, so the rank-normalized full-space model coincides with the full model, and SMOTE plus logistic regression coincides with logistic regression; on PD-252, the full-space kernel models use the archived default G-SMOTE augmentation seed. Partition sensitivity is reported in Section 3.3.

| Dataset | Model | Balanced acc. (%) | Macro-F1 (%) |
| --- | --- | --- | --- |
| UCI-489 | VoxNeuro (full model) | 87.5 ± 4.0 | 87.3 ± 4.0 |
| UCI-489 | Z-standardized full-space model | 85.0 ± 4.2 | 84.7 ± 4.3 |
| UCI-489 | Full-space Euclidean-kernel ablation ($w = 0$) | 86.3 ± 3.6 | 86.0 ± 3.7 |
| UCI-489 | PLS-DA | 82.5 ± 3.1 | 82.2 ± 3.1 |
| UCI-489 | Class-weighted random forest | 80.0 ± 5.0 | 79.9 ± 5.0 |
| UCI-489 | Class-weighted logistic regression | 80.0 ± 2.3 | 79.6 ± 2.6 |
| UCI-489 | Class-weighted linear SVM | 80.0 ± 1.2 | 79.7 ± 1.3 |
| PD-252 | VoxNeuro (full model) | 81.6 ± 4.0 | 81.1 ± 3.4 |
| PD-252 | Rank-normalized full-space model (no subspace stage; G-SMOTE) | 78.2 ± 3.5 | 78.1 ± 2.8 |
| PD-252 | Z-standardized full-space model (G-SMOTE) | 73.1 ± 3.2 | 74.9 ± 3.1 |
| PD-252 | Full-space Euclidean-kernel ablation ($w = 0$; G-SMOTE) | 76.6 ± 3.5 | 75.7 ± 2.8 |
| PD-252 | PLS-DA | 74.7 ± 5.1 | 74.4 ± 5.0 |
| PD-252 | Class-weighted random forest | 67.1 ± 3.7 | 69.6 ± 4.2 |
| PD-252 | Class-weighted logistic regression | 76.9 ± 2.7 | 77.4 ± 2.3 |
| PD-252 | Class-weighted linear SVM | 73.4 ± 4.3 | 74.7 ± 4.3 |
| PD-252 | SMOTE + logistic regression | 77.7 ± 2.8 | 78.6 ± 2.5 |

**Figure 6.**
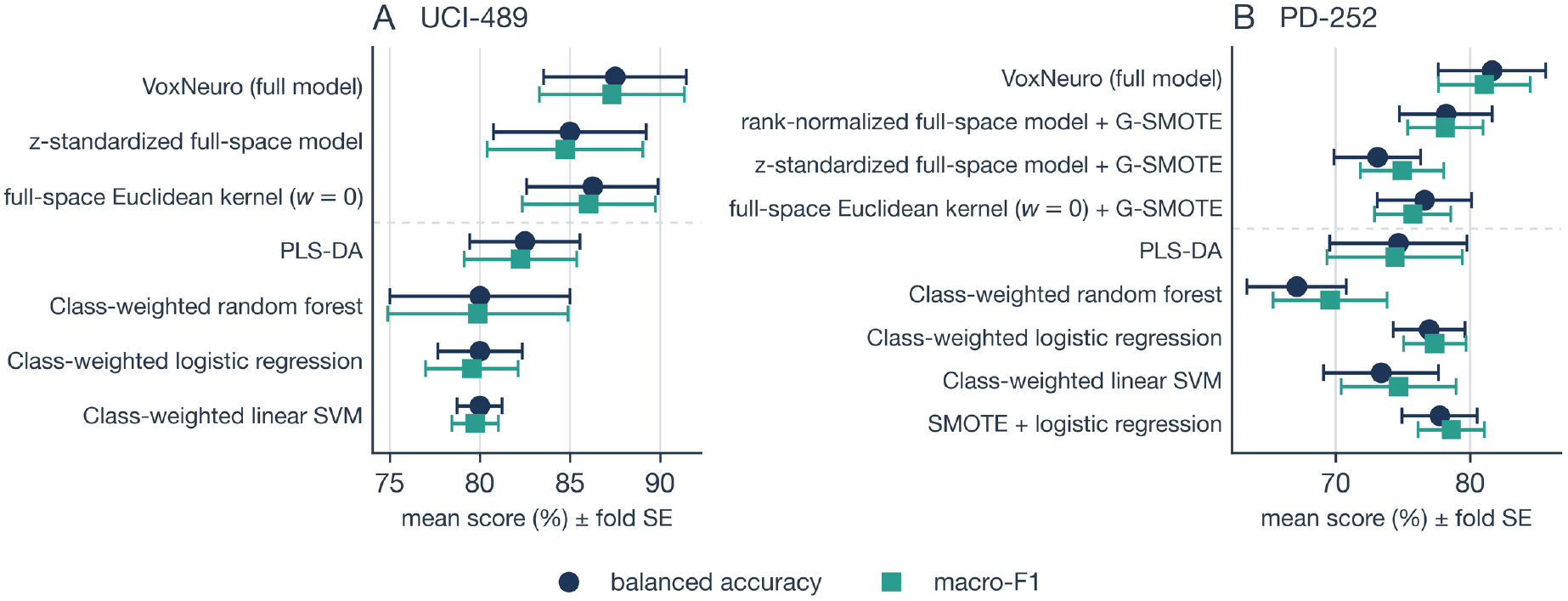
Subject-level scores at the reference partition for (**A**) class-balanced UCI-489 and (**B**) high-dimensional, class-imbalanced PD-252, drawn from the archived out-of-fold results. The full model and its ablations are shown above the dashed separator and the standard comparators below it, all under the same training-fold rank normalization except the z-standardized full-space model. Markers denote mean balanced accuracy (circles) and macro-F1 (squares), in percent; whiskers denote the fold-level standard error across the five outer folds. On UCI-489 the subspace stage is inactive, so the rank-normalized full-space model coincides with the full model and is not shown, and SMOTE plus logistic regression coincides with logistic regression; on PD-252 the full-space Euclidean-kernel ablation retains the G-SMOTE branch.

### 3.2. Operating Characteristics

On UCI-489, VoxNeuro produced 36 true-negative (control), 4 false-positive, 6 false-negative, and 34 true-positive (PD) classifications, yielding sensitivity 85.0%, specificity 90.0%, positive predictive value (PPV) 89.5%, and negative predictive value (NPV) 85.7%. On PD-252, the pooled confusion matrix contained 169 true-positive, 19 false-negative, 47 true-negative, and 17 false-positive classifications, corresponding to sensitivity 89.9%, specificity 73.4%, PPV 90.9%, and NPV 71.2% (Figure 7, Table 5). At the default threshold on PD-252, VoxNeuro had higher specificity than the z-standardized full-space model at its archived default augmentation seed (73.4% versus 53.1%) and lower sensitivity (89.9% versus 93.1%). Table 5 reports operating characteristics computed from pooled subject-level confusion counts, whereas Table 4 reports arithmetic means of the fold-level metrics.

**Table 5.**
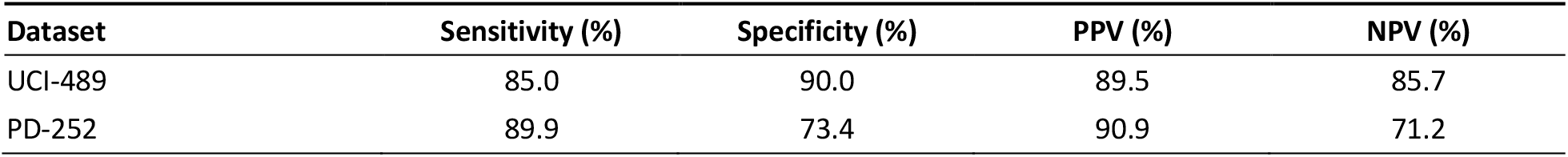
Pooled out-of-fold operating characteristics of VoxNeuro, in percent, computed from the confusion counts of Figure 7 (UCI-489: TP = 34, FN = 6, TN = 36, FP = 4; PD-252: TP = 169, FN = 19, TN = 47, FP = 17). TP, true positive; FN, false negative; TN, true negative; FP, false positive; PPV, positive predictive value; NPV, negative predictive value.

**Figure 7.**
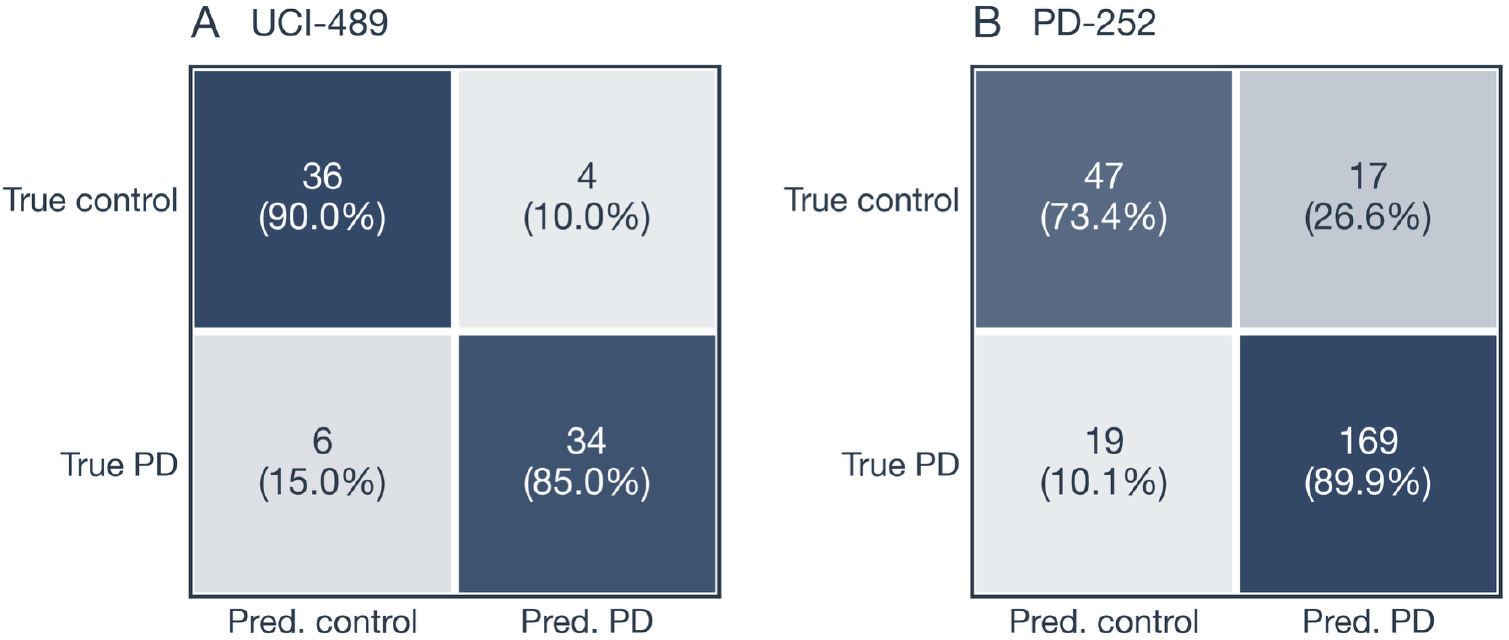
Pooled out-of-fold confusion matrices of VoxNeuro for (**A**) UCI-489 and (**B**) PD-252 at the reference partition, with one prediction per subject. Percentages are normalized within each true class.

### 3.3. Partition Sensitivity and Ablations

Across 20 alternative subject partitions, VoxNeuro achieved a mean balanced accuracy of 78.4 ± 1.7% and a macro-F1 of 78.5 ± 1.4% on PD-252 (mean ± standard deviation, SD, across partitions), exceeding the mean scores of every standard comparator (Figure 8, Table 6). Class-weighted logistic regression was the highest-scoring standard comparator on both metrics (75.2± 2.4% and 75.7 ±2.4%); the paired differences against it were +3.2 ± 2.0 and +2.8 ± 1.8 percentage points, positive in 19 of 20 partitions for each metric. Against each of the remaining standard comparators, VoxNeuro was higher in at least 19 of 20 partitions on both metrics, demonstrating that its advantage on PD-252 is robust to the outer partition. Against the full-space Euclidean-kernel ablation (75.2± 1.8% and 74.4 ± 2.1%), the differences were +3.1 ± 1.5 and +4.1 ± 1.9 percentage points, positive in all 20 partitions. Relative to the z-standardized full-space model (72.1 ± 2.0% and 73.4 ± 1.9%), the paired differences were +6.2 ± 1.8 and +5.0 ± 1.7 percentage points, positive in all 20 partitions on both metrics. Because the augmentation seed of that model varies with the partition in these runs, the comparison was repeated against its archived fixed-augmentation partition results, giving +5.9 ± 1.6 and +4.8 ± 1.7 percentage points, again positive in all 20 partitions. The rank-normalized full-space model exceeded the z-standardized full-space model by mean paired differences of +2.8 percentage points in balanced accuracy and +1.7 in macro-F1, with positive differences in 19 and 17 of 20 partitions. The unaugmented supervised-subspace ensemble of VoxNeuro exceeded the rank-normalized, augmented full-space model by +3.4 and +3.3 percentage points, with positive differences in 19 and 20 of 20 partitions.

**Table 6.** Partition sensitivity and ablations of VoxNeuro (Section 2.11). Each row pairs two models within the same outer subject partition (20 partitions, StratifiedKFold seeds 0–19) or, for the last analysis, within the same augmentation seed at the reference partition. For models with the G-SMOTE branch the augmentation seed is derived from the partition seed, so partition and augmentation realization vary jointly. The Model A and Model B columns report means ± standard deviations across partitions or seeds, in percent; Δ is the paired difference (model A minus model B) in percentage points (pp), and the last column counts strictly positive differences (ties noted where they occur). On UCI-489 the subspace stage is inactive, so VoxNeuro coincides with the rank-normalized full-space model and the normalization comparison is listed once; the forced-gate row applies the subspace stage there regardless. Differences smaller than one percentage point in magnitude are given with two decimals. The subspace branch generates no synthetic subjects, so model B of the last analysis is fixed. Comparisons with the remaining standard comparators are archived with the code.

| Analysis (A vs. B) | Cohort | Metric | Model A (%) | Model B (%) | $\Delta$ (A – B), pp | $\Delta > 0$ |
| --- | --- | --- | --- | --- | --- | --- |
| VoxNeuro vs. z-standardized full-space model | UCI-489 | BA | 85.3 ± 1.9 | 85.3 ± 1.3 | +0.06 ± 2.13 | 9/20 (4 ties) |
| | UCI-489 | macro-F1 | 85.2 ± 1.9 | 85.1 ± 1.4 | +0.06 ± 2.22 | 11/20 |
| | PD-252 | BA | 78.4 ± 1.7 | 72.1 ± 2.0 | +6.2 ± 1.8 | 20/20 |
| | PD-252 | macro-F1 | 78.5 ± 1.4 | 73.4 ± 1.9 | +5.0 ± 1.7 | 20/20 |
| VoxNeuro vs. rank-normalized full-space model | PD-252 | BA | 78.4 ± 1.7 | 75.0 ± 1.6 | +3.4 ± 1.7 | 19/20 |
| | PD-252 | macro-F1 | 78.5 ± 1.4 | 75.1 ± 1.8 | +3.3 ± 1.7 | 20/20 |
| Full-space model: rank normalization vs. z-standardization | PD-252 | BA | 75.0 ± 1.6 | 72.1 ± 2.0 | +2.8 ± 2.3 | 19/20 |
| | PD-252 | macro-F1 | 75.1 ± 1.8 | 73.4 ± 1.9 | +1.7 ± 2.3 | 17/20 |
| VoxNeuro vs. full-space Euclidean-kernel ablation ($w = 0$) | UCI-489 | BA | 85.3 ± 1.9 | 85.1 ± 2.0 | +0.25 ± 2.58 | 9/20 (2 ties) |
| | UCI-489 | macro-F1 | 85.2 ± 1.9 | 84.9 ± 2.1 | +0.26 ± 2.62 | 10/20 (1 tie) |
| | PD-252 | BA | 78.4 ± 1.7 | 75.2 ± 1.8 | +3.1 ± 1.5 | 20/20 |
| | PD-252 | macro-F1 | 78.5 ± 1.4 | 74.4 ± 2.1 | +4.1 ± 1.9 | 20/20 |
| VoxNeuro vs. class-weighted logistic regression | UCI-489 | BA | 85.3 ± 1.9 | 82.8 ± 2.7 | +2.5 ± 3.0 | 16/20 (1 tie) |
| | UCI-489 | macro-F1 | 85.2 ± 1.9 | 82.6 ± 2.7 | +2.6 ± 3.0 | 17/20 |
| | PD-252 | BA | 78.4 ± 1.7 | 75.2 ± 2.4 | +3.2 ± 2.0 | 19/20 |
| | PD-252 | macro-F1 | 78.5 ± 1.4 | 75.7 ± 2.4 | +2.8 ± 1.8 | 19/20 |
| VoxNeuro vs. PLS-DA | UCI-489 | BA | 85.3 ± 1.9 | 83.1 ± 2.4 | +2.2 ± 3.4 | 12/20 (3 ties) |
| | UCI-489 | macro-F1 | 85.2 ± 1.9 | 82.9 ± 2.5 | +2.3 ± 3.5 | 13/20 (1 tie) |
| | PD-252 | BA | 78.4 ± 1.7 | 74.2 ± 2.3 | +4.1 ± 2.2 | 19/20 |
| | PD-252 | macro-F1 | 78.5 ± 1.4 | 74.1 ± 2.1 | +4.3 ± 2.0 | 20/20 |
| Subspace stage forced on vs. gate rule | UCI-489 | BA | 84.2 ± 2.2 | 85.3 ± 1.9 | –1.1 ± 2.1 | 4/20 (4 ties) |
| | UCI-489 | macro-F1 | 84.0 ± 2.2 | 85.2 ± 1.9 | –1.1 ± 2.2 | 6/20 (1 tie) |
| VoxNeuro vs. VoxNeuro without the gender covariate | UCI-489 | BA | 85.3 ± 1.9 | 83.8 ± 1.7 | +1.5 ± 2.6 | 13/20 (3 ties) |
| | UCI-489 | macro-F1 | 85.2 ± 1.9 | 83.7 ± 1.8 | +1.5 ± 2.7 | 14/20 |
| | PD-252 | BA | 78.4 ± 1.7 | 77.0 ± 1.6 | +1.4 ± 1.3 | 19/20 |
| | PD-252 | macro-F1 | 78.5 ± 1.4 | 77.1 ± 1.5 | +1.3 ± 1.4 | 18/20 |
| Subspace branch with G-SMOTE vs. without (24 augmentation seeds) | PD-252 | BA | 79.7 ± 0.8 | 81.6 (fixed) | –1.9 ± 0.8 | 1/24 |
| | PD-252 | macro-F1 | 80.4 ± 0.8 | 81.1 (fixed) | –0.65 ± 0.85 | 7/24 |

**Figure 8.**
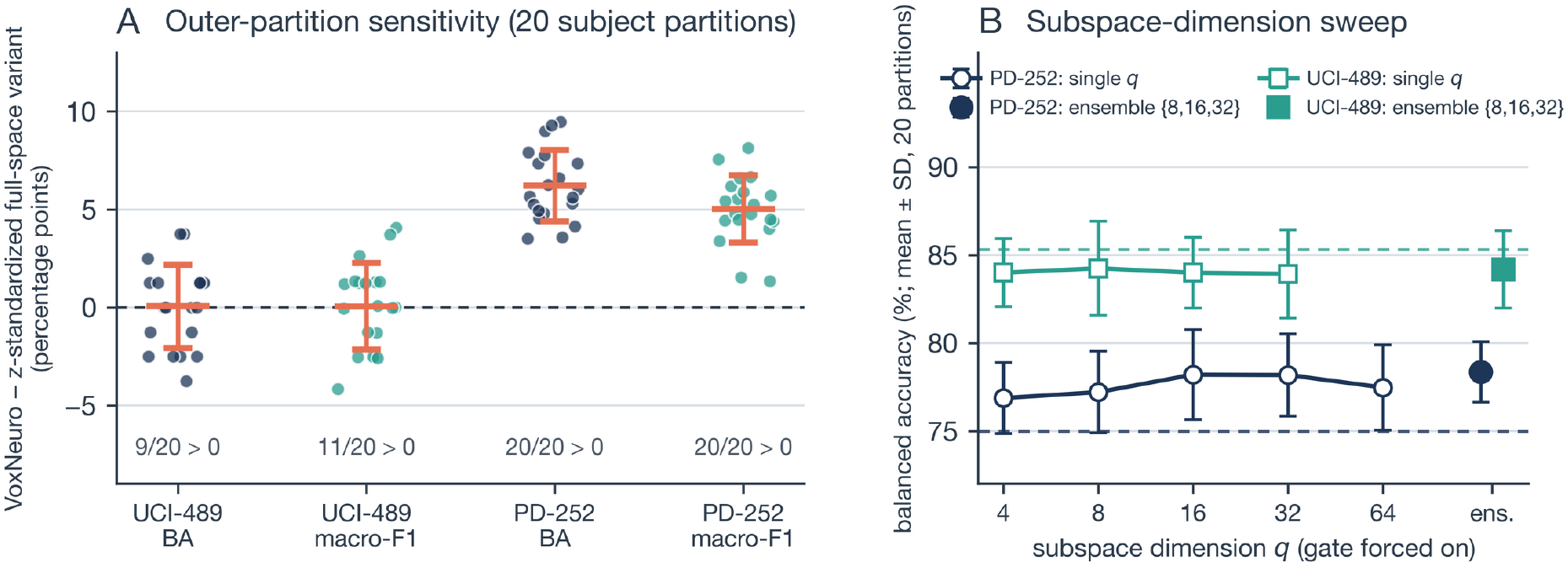
Sensitivity of VoxNeuro to the outer subject partition and to the subspace dimension. (**A**) Paired differences, in percentage points, between VoxNeuro and the z-standardized full-space model across 20 alternative outer subject partitions (points; coral bars denote mean ± SD; counts give the number of partitions with a positive difference), for balanced accuracy (BA, navy) and macro-F1 (teal) on both cohorts. On PD-252, the z-standardized full-space model’s augmentation seed varies with the partition. On UCI-489, augmentation and the subspace stage are inactive, and the comparison evaluates rank normalization against z-standardization. (**B**) Subspace-dimension sweep with the gate forced on: mean balanced accuracy (%) across the 20 partitions (± SD) for single members *q* ∈ {4,8,16,32,64} (*q* ≤ 32 on UCI-489), the three-member ensemble (filled markers), and the rank-normalized full-space model of each cohort (dashed horizontal line).

On UCI-489, where the subspace stage is inactive, VoxNeuro averaged 85.3 ± 1.9% and 85.2 ± 1.9%. It exceeded class-weighted logistic regression in 16 of 20 partitions and PLS-DA in 12 of 20 in balanced accuracy (Table 6). Its scores were similar to those of the full-space Euclidean-kernel ablation (mean paired differences +0.25 and +0.26 percentage points; VoxNeuro higher in 9 and 10 of 20 partitions, with 2 and 1 ties). They were also similar to those of the z-standardized full-space model (85.3 ± 1.3% and 85.1 ± 1.4%; paired balanced-accuracy difference +0.06 ± 2.13 percentage points). Forcing the subspace stage on for UCI-489 lowered balanced accuracy by 1.1 percentage points on average (higher in 4 of 20 partitions), consistent with the gate rule.

At the reference partition on PD-252, the rank-normalized full-space model reached 78.2% and 78.1% and the z-standardized full-space model 73.1% and 74.9%, so rank normalization improved balanced accuracy by 5.1 percentage points and macro-F1 by 3.2; replacing the augmented full-space model with the unaugmented supervised-subspace ensemble improved them by a further 3.4 and 2.9 percentage points. The three ensemble members individually reached 80.9%, 81.4%, and 79.8% balanced accuracy for *q* = 8, 16, and 32, and the ensemble exceeded each of them. Across the five reference-partition training folds, the mean coefficient of variation of the full-space chordal distances on PD-252 was 0.019 under z-standardization and 0.035 under rank normalization; the corresponding values on UCI-489 were 0.052 and 0.066, and for the PD-252 representation projected to 16 dimensions the coefficient of variation was 0.064.

Figure 8B shows the subspace-dimension sweep with the gate forced on for both cohorts. On PD-252, single members reached 76.9% (*q* = 4), 77.2% (*q* = 8), 78.2% (*q* = 16), 78.2% (*q* = 32), and 77.5% (*q* = 64) mean balanced accuracy across partitions, and the three-member ensemble achieved the highest mean balanced accuracy among these tested configurations, at 78.4%. On UCI-489, mean balanced accuracy ranged from 83.9% to 84.3% across the tested subspace dimensions, below the full-space model’s 85.3%. Inside the subspace branch, geodesic oversampling to class parity changed balanced accuracy by −1.9 ± 0.8 percentage points and macro-F1 by −0.65 ± 0.85 across the 24 augmentation seeds (positive in 1 and 7 seeds; Table 6). The unaugmented class-weighted ensemble achieved higher mean balanced accuracy and macro-F1 in this ablation. The view ablations retained the automatic gate, so they evaluate the supervised-subspace ensemble on PD-252 and the rank-normalized full-space model on UCI-489. Across partitions, the Grassmann-only variants (*w* = 1) reached balanced accuracies of 63.6% and 57.9%, respectively, and the Euclidean-only variants (*w* = 0) reached 77.9% and 85.1%. Adding the Grassmann kernel increased mean balanced accuracy over the matched Euclidean-only variant by 0.48 percentage points on PD-252 and 0.25 on UCI-489. The differences were positive in 12 of 20 PD-252partitions and 9 of 20 UCI-489 partitions, with two ties on UCI-489.

In the covariate ablation, we removed the dataset-provided gender variable from the feature set of VoxNeuro, the five standard comparators, and the rank-normalized full-space kernel models and refitted every pipeline without it. At the reference partition, the balanced accuracy and macro-F1 of VoxNeuro changed from 87.5% and 87.3% to 83.8% and 83.5% on UCI-489 and from 81.6% and 81.1% to 79.8% and 80.1% on PD-252. Across the 20 partitions, the paired decreases were 1.5 ± 2.6 and 1.5 ± 2.7 percentage points on UCI-489 (lower without the covariate in 13 and 14 of 20 partitions, with 3 ties in balanced accuracy). On PD-252 they were 1.4 ± 1.3 and 1.3 ± 1.4 percentage points (lower in 19 and 18 of 20; Table 6). The mean balanced accuracy of the standard comparators changed by less than 0.5 percentage points on PD-252 and by up to 3.6 percentage points (class-weighted linear SVM) on UCI-489. VoxNeuro remained above every standard comparator at the reference partition on both cohorts without the covariate, and on PD-252 the number of partitions in which it exceeded a standard comparator ranged from 14 to 20, depending on the comparator. These differences quantify the incremental value of retaining the covariate within the evaluated pipeline; their interpretation is discussed in Section 4.

## 4. Discussion

In this study, we introduced VoxNeuro, a subject-level framework for repeated Parkinsonian speech in which the evaluation unit coincides with the clinical unit. At the reference five-fold subject-disjoint partition, VoxNeuro achieved balanced accuracies of 87.5% on UCI-489 and 81.6% on PD-252 and exceeded all five standard comparators on both balanced accuracy and macro-F1. Across 20 additional PD-252 partitions, it exceeded each standard comparator in at least 19 partitions and the z-standardized full-space model in all 20 for both metrics, with preprocessing fitted within each outer training fold. These results demonstrate that VoxNeuro improves subject-level classification over the standard comparators at the reference partition on both cohorts, and that its advantage on the imbalanced, high-dimensional PD-252 cohort is robust to the choice of subject partition. The Grassmann representation encodes the joint orientation of each subject’s repeated recordings, and its basis invariance removes arbitrary choices of orthonormal basis. The Euclidean view retains coordinate-level location and dispersion.

Our ablations characterize the effects of normalization and branch configuration and quantify the performance differences associated with view fusion and the gender covariate. On PD-252, rank normalization improved balanced accuracy by 5.1 percentage points at the reference partition and by 2.8 on average across partitions, and it increased the coefficient of variation of the full-space chordal distances (Section 3.3), indicating greater relative dispersion of those distances. Replacing the augmented full-space model with the unaugmented supervised-subspace ensemble improved balanced accuracy by a further 3.4 percentage points at the reference partition and on average across partitions, and the three-member ensemble achieved the highest mean balanced accuracy among the tested subspace configurations. At the reference partition and default threshold, VoxNeuro had higher specificity than the z-standardized full-space model at its archived default augmentation seed (73.4% versus 53.1%) and lower sensitivity (89.9% versus 93.1%). The branch choice tracks cohort dimensionality: on the 45-dimensional UCI-489 cohort, forcing the supervised-subspace stage reduced mean balanced accuracy by 1.1 percentage points, consistent with retaining the full-space representation under the dimensionality rule. In the matched view ablations, adding the Grassmann kernel to the Euclidean configuration increased mean balanced accuracy by 0.48 percentage points on PD-252 and 0.25 on UCI-489. Retaining the dataset-provided gender covariate increased mean balanced accuracy across partitions by 1.5 percentage points on UCI-489 and 1.4 on PD-252; after its removal and refitting of all pipelines, VoxNeuro remained above every standard comparator at the reference partition on both cohorts. Together, these comparisons show consistent improvements on the high-dimensional cohort from the two fold-local elements introduced here: rank normalization within the full-space model, and the replacement of the augmented full-space model by the unaugmented supervised-subspace ensemble.

The risk decomposition in Equations (19) and (20) explains why subject identity must be considered when evaluating repeated recordings. When seen-person risk is lower than unseen-person risk, record-wise splitting produces an optimistic estimate whose magnitude depends on the probability of subject overlap. Subject-disjoint folds remove this overlap term and align evaluation with prediction for previously unseen participants [20,22].

Evaluation design is central to interpreting voice-based PD classification results [16,17]. VoxNeuro evaluates one prediction per previously unseen participant, following the use-case alignment described by Saeb et al. [20], and its treatment of repeated recordings relates to the replication-aware classification developed for UCI-489 [29]. The present framework represents each subject through subspace geometry and Euclidean summaries and compares the resulting configurations under shared subject partitions and preprocessing. These subject-disjoint estimates provide a reference for future methods evaluated under the same protocol on these corpora.

A research-stage web interface illustrates browser-based capture with a commodity microphone for sustained vowels, a read passage using an excerpt of the Rainbow Passage [69], and free speech (Figure 9). The interface also includes a longitudinal display with illustrative session entries. Figure 10 relates the capture workflow to the subject-level analysis framework. Repeated speech collection of this kind is consistent with technology roadmaps for PD care [50,51].

**Figure 9.**
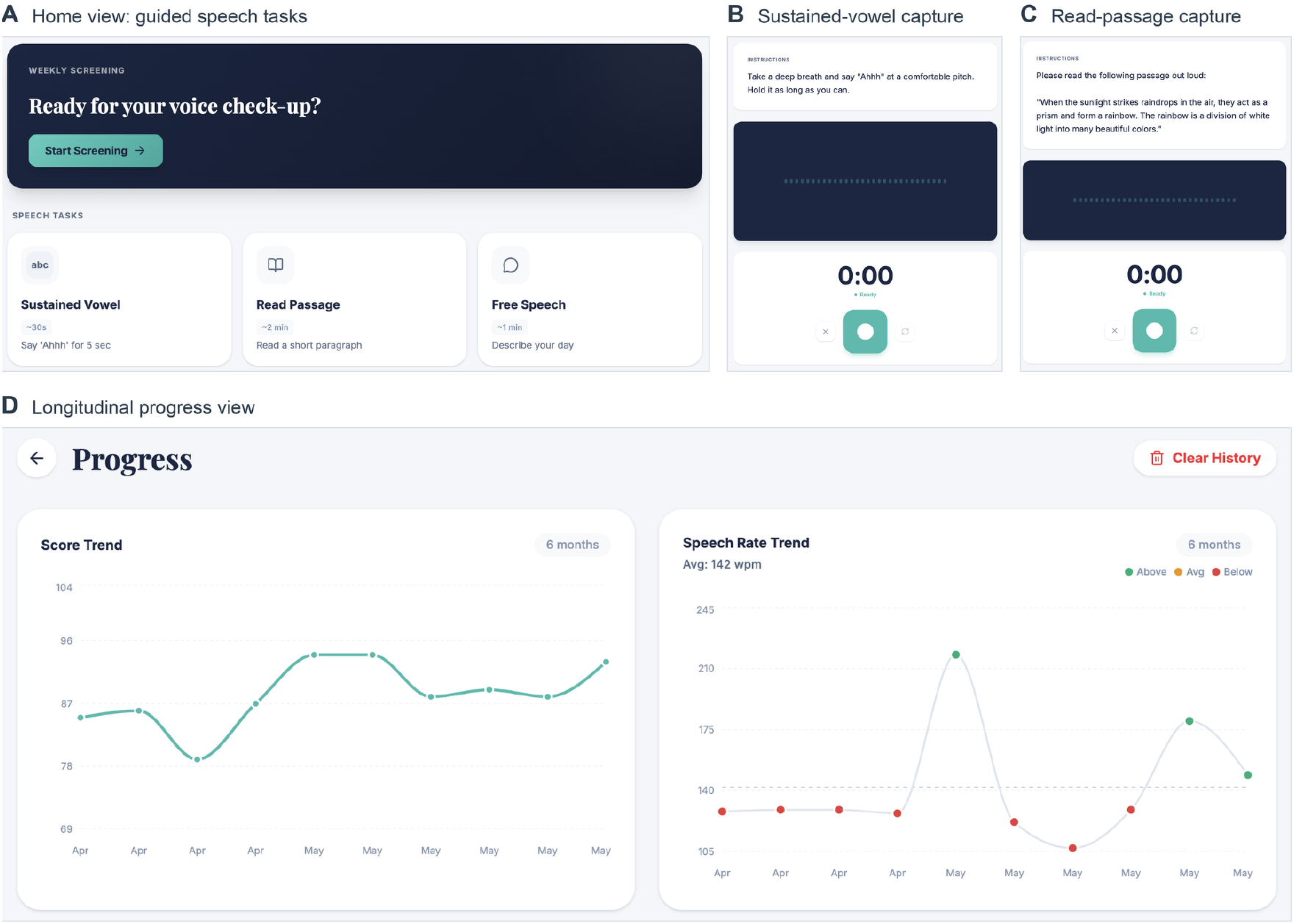
VoxNeuro web application (research-stage prototype, reachable at https://voxneuro.org). (**A**) Home view listing the three guided speech tasks. (**B**) Sustained-vowel capture screen. (**C**) Read-passage capture screen showing an excerpt from the Rainbow Passage. (**D**) Longitudinal progress view with illustrative entries. The application requires only a commodity microphone and a web browser; no application-collected data were used in the analyses reported here.

**Figure 10.**
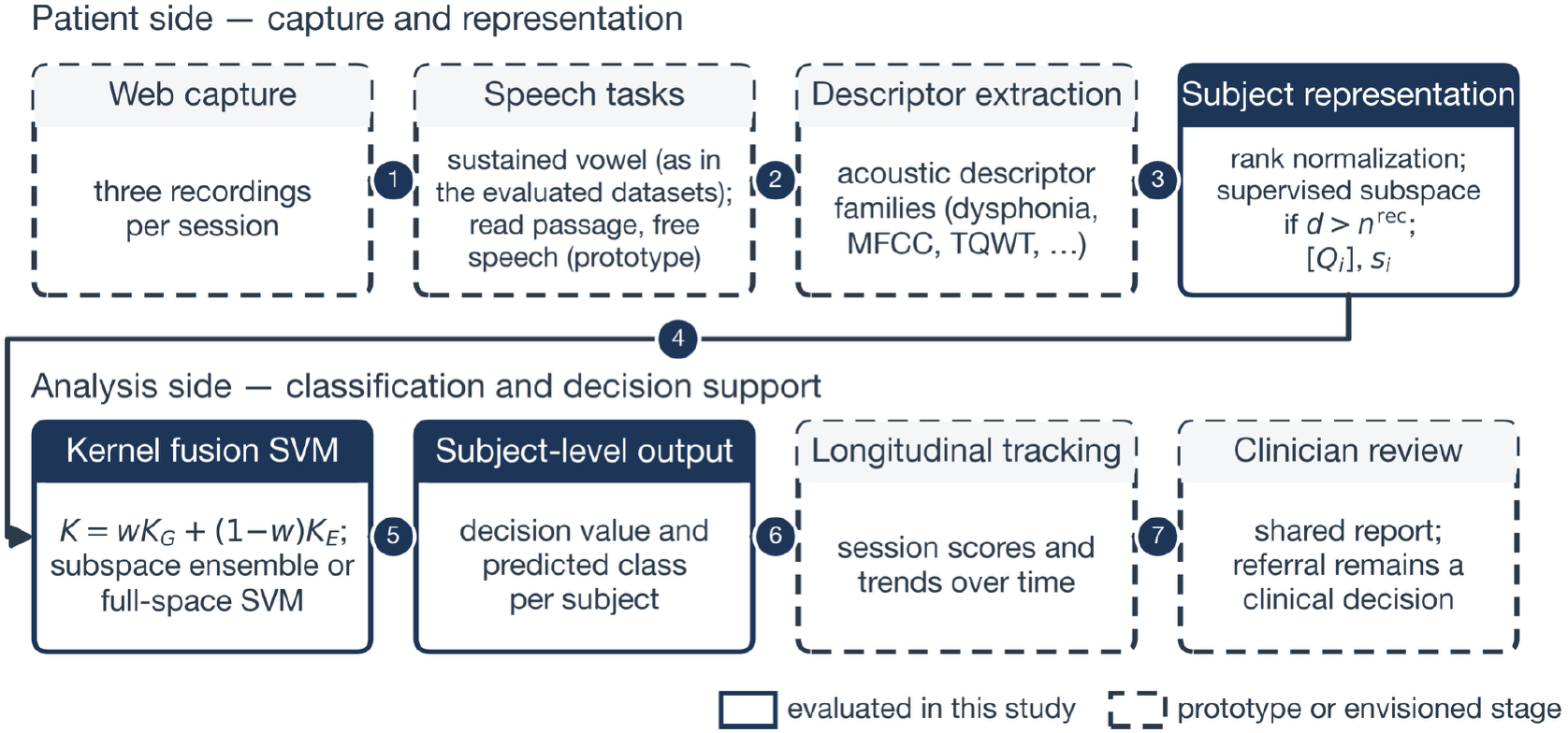
Study pipeline and envisioned deployment workflow of the VoxNeuro prototype. Solid boxes mark the components evaluated offline in this study on the two retrospective public cohorts (rank normalization and subject representation, with the supervised-subspace branch in the high-dimensional regime and the full-space branch with conditional G-SMOTE otherwise; kernel fusion; and subject-level scoring). Dashed boxes mark the capture and decision-support stages of the prototype, which were not part of the evaluation. Descriptor extraction in this study used the dataset-provided engineered features, and any referral decision remains a clinical judgment. The evaluated cohorts consist of three sustained-vowel recordings per subject from the public datasets; the read-passage and free-speech tasks are prototype capture tasks.

This study has limitations. The shared configuration was developed on the reference partition of both cohorts, making the performance estimates susceptible to method-selection optimism [23,24]. The 20-partition analysis assesses sensitivity within the same 80 and 252 subjects and does not constitute external validation. Fold-level standard errors describe variability among dependent folds, and no inferential test of model superiority was performed [25]. Evaluation used dataset-provided engineered features and five standard comparators; pretrained audio models, other deep architectures, probability calibration [70,71], decision-curve analysis [72], and device, language, and prespecified subgroup evaluations were outside its scope. Application-collected speech, heterogeneous-task representations, and longitudinal clinical performance were not evaluated. PPV and NPV depend on cohort class composition and the decision threshold and therefore cannot be transferred directly to a screening population; the PD-252 specificity corresponds to positive classifications for roughly one in four controls. The full-space and supervised-subspace comparison jointly changes projection, ensembling, and oversampling, while the subspace-oversampling ablation also changes the training sample used to scale ensemble decisions, so these comparisons do not isolate the individual effects of those components. The Grassmann term produced small mean improvements over the matched Euclidean-only configurations, and its standalone performance was substantially lower. For the two rank-deficient subjects, the third SVD direction is a numerical completion, and the rank-two sensitivity analysis concerns the z-standardized full-space variant. Geodesic oversampling was inactive in the complete-model evaluations, retains potential noise-amplification risks [47], and does not establish that one set of recordings can realize both interpolated views. The binary covariate does not distinguish sex from gender, differs in distribution between the diagnostic classes, and receives a comparatively large rank-transformed scale; its removal measures a pipeline-level performance difference without separating demographic from acoustic information. The dimensionality gate was evaluated on two cohorts, and PLS fitting treats each subject’s correlated recordings as separate observations. The quantile-knot rule, subspace dimensions, and ensemble weights were determined during development, and the exploratory analyses of the full-space variant do not establish systematic sensitivity of the final VoxNeuro configuration to the SVM penalty, neighbor count, bandwidth rule, or fusion-weight selection.

Future work will focus on prospective evaluation of repeated raw speech, external evaluation of the frozen model, and extension to additional recording tasks. Preregistered preprocessing and thresholds, together with reporting aligned with clinical prediction guidance [73,74], will support the planned calibration, test–retest, fairness, decision-curve, and pretrained-model analyses. Prospective application use will require privacy, consent, security, accessibility, and clinical governance procedures. Additional task-specific views can be combined through nonnegative sums of positive-semidefinite kernels, with weights selected through nested multiple-kernel optimization [42,43].

## 5. Conclusions

In this study, we presented VoxNeuro, which combines Grassmann representations of repeated recordings with Euclidean summaries through a provably positive-semidefinite kernel. At the reference subject-disjoint partition, VoxNeuro achieved balanced accuracies of 87.5% on UCI-489 and 81.6% on PD-252, exceeding all five standard comparators under identical folds and normalization as well as its full-space ablations. Across 20 additional PD-252 partitions, it exceeded each standard comparator in at least 19 partitions for both balanced accuracy and macro-F1. At the reference partition on PD-252, rank normalization improved the balanced accuracy of the full-space model by 5.1 percentage points, and replacing the augmented full-space model with the unaugmented supervised-subspace ensemble improved it by a further 3.4. Together with the archived predictions and analysis scripts, VoxNeuro provides a reproducible framework for subject-level classification of repeated speech measurements in PD screening research, with a comparative advantage that was robust across the evaluated PD-252 subject partitions. The framework is computationally tractable for cohorts of this size and extends to additional speech tasks through further positive-semidefinite kernel terms.

## Data Availability

The datasets analyzed in this study are openly available from the UCI Machine Learning Repository: the Parkinson Dataset with Replicated Acoustic Features (UCI-489; UCI dataset 489, doi:10.24432/C5701F) and the Parkinson Disease Classification dataset (PD-252; UCI dataset 470, doi:10.24432/C5MS4X).

## Author Contributions

Conceptualization, L.K.; methodology, L.K.; software, L.K.; validation, L.K. and M.W.; formal analysis, L.K.; investigation, L.K. and M.W.; data curation, L.K.; writing—original draft preparation, L.K.; writing—review and editing, L.K. and M.W.; visualization, L.K.; project administration, L.K. All authors have read and agreed to the published version of the manuscript.

## Funding

This research received no external funding.

## Institutional Review Board Statement

Ethical review and approval were not required for this study, as it consisted exclusively of a secondary computational analysis of publicly available, fully de-identified datasets; no participants were recruited, and no new human data were collected.

## Informed Consent Statement

Not applicable. The study analyzed publicly available, de-identified datasets and did not involve direct human participation; informed consent and ethics procedures for the original data collections are documented in the source publications cited in Section 2.1.

## Data Availability Statement

The datasets analyzed in this study are openly available from the UCI Machine Learning Repository: the Parkinson Dataset with Replicated Acoustic Features (UCI-489; UCI dataset 489, doi:10.24432/C5701F) and the Parkinson’s Disease Classification dataset (PD-252; UCI dataset 470, doi:10.24432/C5MS4X). Source code is available at https://github.com/khan-laiba/voxneuro (release v2.1.9).

## Conflicts of Interest

The authors declare no conflicts of interest.

## Appendix A.

**Supplementary Analyses of a z-Standardized Full-Space Variant**

These supplementary analyses assess the sensitivity of the z-standardized full-space configuration, comprising the fused kernel with *w* = 0.5 and conditional G-SMOTE, under the variants specified below. All analyses used the protocol of Section 2.11 and are archived with the reference implementation.

### Appendix A.1. Sensitivity to the Augmentation Seed

On PD-252, G-SMOTE randomly samples anchor subjects and neighbors and draws interpolation coefficients, so performance varies with the augmentation seed. To isolate this source of variability, the fixed five-fold evaluation was repeated for 24 distinct augmentation seeds while the outer-fold assignments, preprocessing, hyperparameters, and all non-augmentation settings were held fixed. Within each seed and fold, the fused model and the matched Euclidean-kernel ablation used the same augmented training set, so their difference is paired at the seed level. The reference-partition results of the full-space kernel models in Table 4 use the reference implementation’s default augmentation seed, which is archived and regenerable; this appendix characterizes augmentation-seed variability around it for the z-standardized full-space model.

The paired analysis characterizes the sensitivity of the z-standardized full-space model, of its Euclidean-kernel ablation, and of their difference to the augmentation seed (Table A1); VoxNeuro generated no synthetic subjects on either evaluated cohort and is unaffected by this seed. Across 24 seed runs, balanced accuracy was 73.9 ± 1.2% for the fused model and 73.1 ± 1.3% for the matched Euclidean-kernel ablation, and macro-F1 was 75.2 ± 1.3% and 72.9 ± 1.3%, respectively (mean ± standard deviation across seeds). The paired macro-F1 difference averaged +2.3 ± 1.5 percentage points and was positive in 23 of 24 runs. The paired balanced-accuracy difference averaged +0.86 ± 1.35 percentage points, was positive in 16 of 24 runs, and reversed sign in 8 of 24. The archived default-seed realization of Table 4 yielded balanced accuracy of 73.1% and macro-F1 of 74.9%, within the respective seed ranges of 72.2% to 76.0% and 73.2% to 77.7%. Across the same 24 seeds, the fully Euclidean pipeline (ordinary SMOTE followed by the Euclidean radial-basis-function (RBF) kernel) averaged 72.6 ± 0.9% and 71.8 ± 1.0%. Replacing ordinary SMOTE by G-SMOTE in the Euclidean-kernel model changed balanced accuracy by +0.51 ± 1.55 and macro-F1 by +1.1 ± 1.6 percentage points (positive in 15 and 17 of 24 seeds). The fused G-SMOTE pipeline exceeded the fully Euclidean pipeline by +1.4 ± 1.5 and +3.4 ± 1.6 percentage points (positive in 19 and 24 of 24 seeds). Thus, conditional on the fixed outer folds, the macro-F1 margin of the fused kernel over its Euclidean ablation was directionally consistent in all but one seed run, whereas the balanced-accuracy margin can be reversed by individual draws.

**Table A1.**
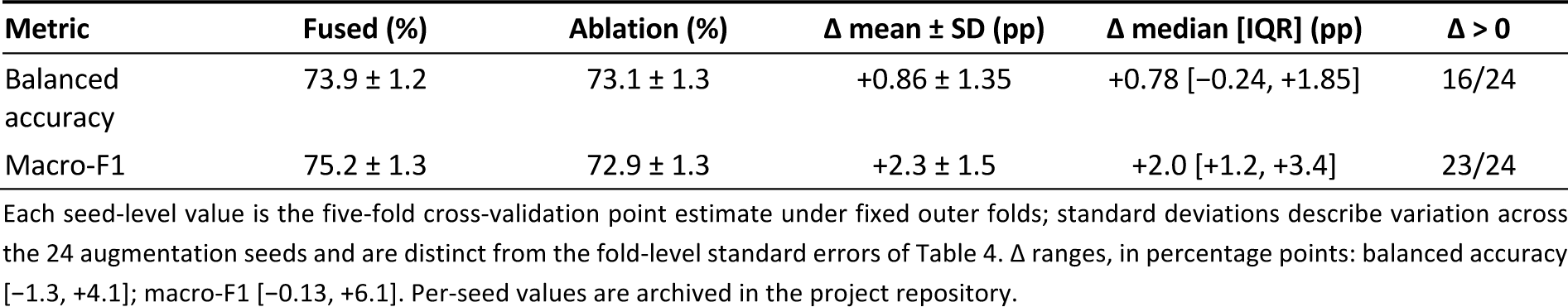
Sensitivity of the full-space branch to the G-SMOTE augmentation seed on PD-252 (z-standardized full-space model versus its Euclidean-kernel ablation; Appendix A.1): paired evaluation across 24 distinct seeds with outer folds, preprocessing, and hyperparameters held fixed. The Fused and Ablation columns report means ± standard deviations (SD) across seeds, in percent; Δ denotes the paired difference (fused minus ablation) in percentage points (pp), computed within each seed, summarized by its mean ± SD, median [interquartile range, IQR], and sign count.

| Metric | Fused (%) | Ablation (%) | $\Delta$ mean ± SD (pp) | $\Delta$ median [IQR] (pp) | $\Delta > 0$ |
| --- | --- | --- | --- | --- | --- |
| Balanced accuracy | $73.9 \pm 1.2$ | $73.1 \pm 1.3$ | $+0.86 \pm 1.35$ | $+0.78 [-0.24, +1.85]$ | 16/24 |
| Macro-F1 | $75.2 \pm 1.3$ | $72.9 \pm 1.3$ | $+2.3 \pm 1.5$ | $+2.0 [+1.2, +3.4]$ | 23/24 |

### Appendix A.2. Feature-Family Permutation Sensitivity

For feature family *g*, the fold-level decrease in performance and its summary are

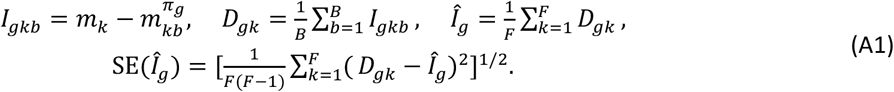

Sensitivity of the Euclidean summary of the z-standardized full-space model was evaluated on each held-out fold without refitting. The coordinates of *s*_*i*_ associated with family *g* were jointly permuted across held-out subjects while *Q*_*i*_ and all other summary coordinates were held fixed. In Equation (A1), *m*_*k*_ is the original held-out macro-F1 and 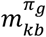 is macro-F1 after the *b*-th permutation draw. Per family and fold, *B* = 20 permutation draws were made with a fixed random seed. The decrease was averaged over draws before the fold-level summary, and the six families follow the descriptor taxonomy of the source dataset (column assignments are archived with the code).

The TQWT summary block produced the largest mean decrease in held-out macro-F1, at 14.9 ± 2.0 percentage points, followed by the MFCC summary block, at 4.6 ± 0.8 percentage points (mean ± fold-level SE; Figure A1). Within a fold, the standard deviation across the 20 permutation draws averaged 6.7 percentage points for TQWT. The vocal/time-frequency, wavelet, IMF/EMD, and demographic families produced mean decreases near zero. These values measure the dependence of the fitted model on the Euclidean summary coordinates of each family under correlated features; the permutation leaves the Grassmann view unchanged. The prominence of the TQWT family is consistent with the descriptor engineering of the source dataset, where tunable Q-factor features were introduced precisely for Parkinsonian voice [31,55].

**Figure A1.**
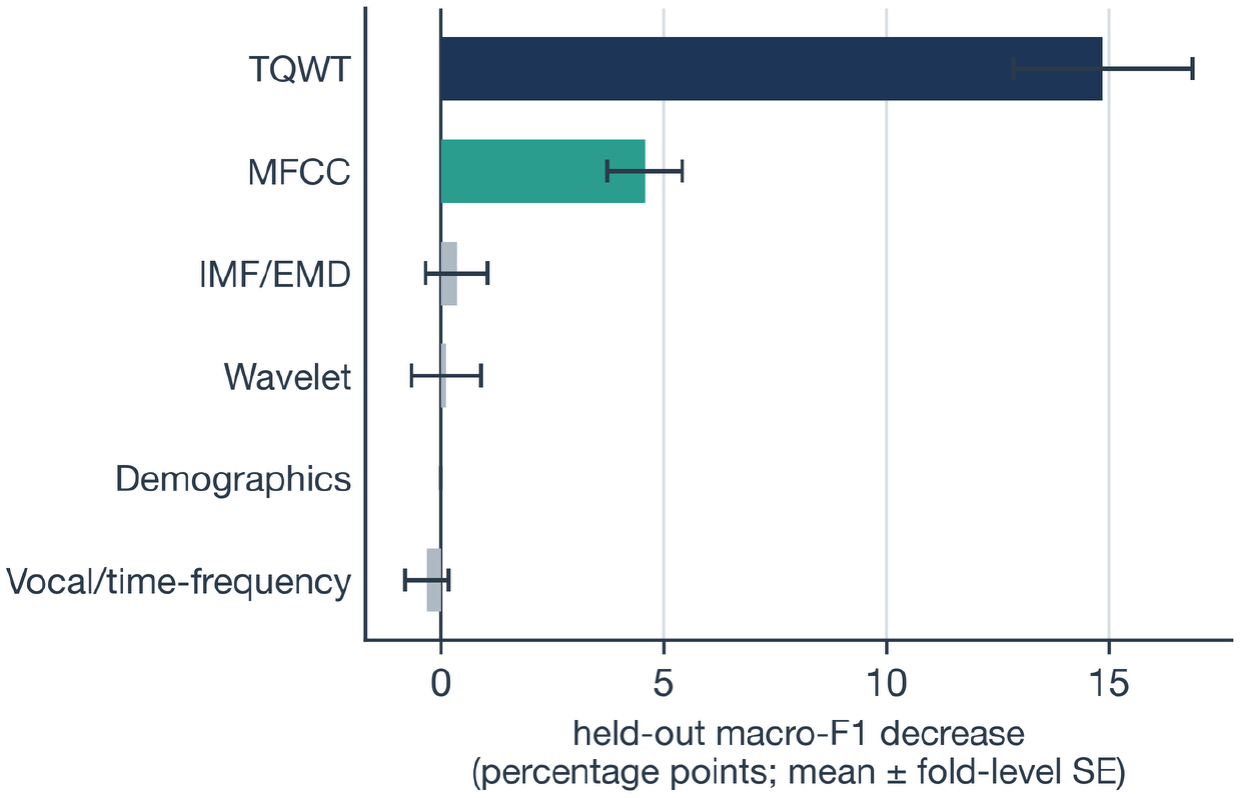
PD-252 feature-family permutation sensitivity of the Euclidean summary view of the z-standardized full-space model (Appendix A.2) under the archived default-seed realization (20 permutation draws per family and fold). Positive values denote a decrease in held-out macro-F1, in percentage points, after permutation, and whiskers denote fold-level standard error. TQWT denotes tunable Q-factor wavelet transform; MFCC denotes Mel-frequency cepstral coefficients; IMF/EMD denotes intrinsic mode function/empirical mode decomposition; Demographics denotes the dataset-provided gender covariate.

### Appendix A.3. Partition, Rank, Augmentation-Family, and Fusion-Weight Sensitivity

Four supplementary analyses of the z-standardized full-space model were performed with the protocol of Section 2.11. First, the five-fold evaluation was repeated for 20 alternative stratified subject partitions (StratifiedKFold seeds 0–19) with a fixed augmentation seed, and paired differences between models were summarized across partitions. Second, the fused model was rerun retaining *r* = 2 singular vectors, a rank supported by every subject’s recording matrix, to assess sensitivity to the retained rank and to the completion directions used for the two rank-deficient subjects noted in Section 2.3. Third, on PD-252 the augmentation family was varied across the same 24 seeds as Appendix A.1, so that the fused model, the matched Euclidean-kernel ablation with G-SMOTE, and the fully Euclidean pipeline were compared with paired draws. Fourth, the fixed-weight sweep evaluated *w* ∈ {0,0.1, …,1.0} in every seed and partition run (Figure A2 displays 0 ≤ *w* ≤ 0.8). A nested variant selected *w* ∈ {0,0.1, …,0.8} inside each training fold by repeated inner cross-validation comprising three repetitions of five folds, with balanced accuracy as the selection metric, so that the outer held-out folds were never used for selection. The inner selection used representations normalized on the complete outer-training fold.

Across the 20 partitions, the z-standardized full-space model and its matched Euclidean-kernel ablation had similar mean estimates on both cohorts (paired balanced-accuracy differences +0.44 ± 1.30 percentage points on UCI-489 and −0.05 ± 1.63 on PD-252; macro-F1 +0.46 ± 1.31 and +0.77 ± 1.75). The fused model retained a macro-F1 advantage of +2.8 ± 1.9 percentage points over the fully Euclidean pipeline on PD-252 (18 of 20 partitions). With *r* = 2, the z-standardized full-space model achieved balanced accuracy and macro-F1 of 83.8% and 83.4% on UCI-489 and of 73.9% and 75.5% on PD-252. Figure A2 shows the sensitivity of the full-space model to the fusion weight. Across the 24 PD-252 augmentation seeds, weights *w* = 0.2–0.7 exceeded the *w* = 0 ablation in macro-F1 in 22 or 23 runs, whereas *w* = 0.1 did so in 15 runs. The highest mean balanced accuracy occurred at *w* = 0.3 and the highest mean macro-F1 at *w* = 0.7, and on UCI-489 the highest observed means also occurred at larger weights (*w* = 0.7). Nested selection of the weight inside each training fold produced positive balanced-accuracy differences against the Euclidean-kernel ablation in 20 of 24 PD-252 seed runs and 13 of 20 PD-252 partition runs, with mean differences of +0.55 and +0.07 percentage points. Additional full-space variants (dimension-reduced, rank-one, centered, and block-balanced Grassmann views, decision-level fusion, weight ensembles, and other penalties) are documented in the project repository.

**Figure A2.**
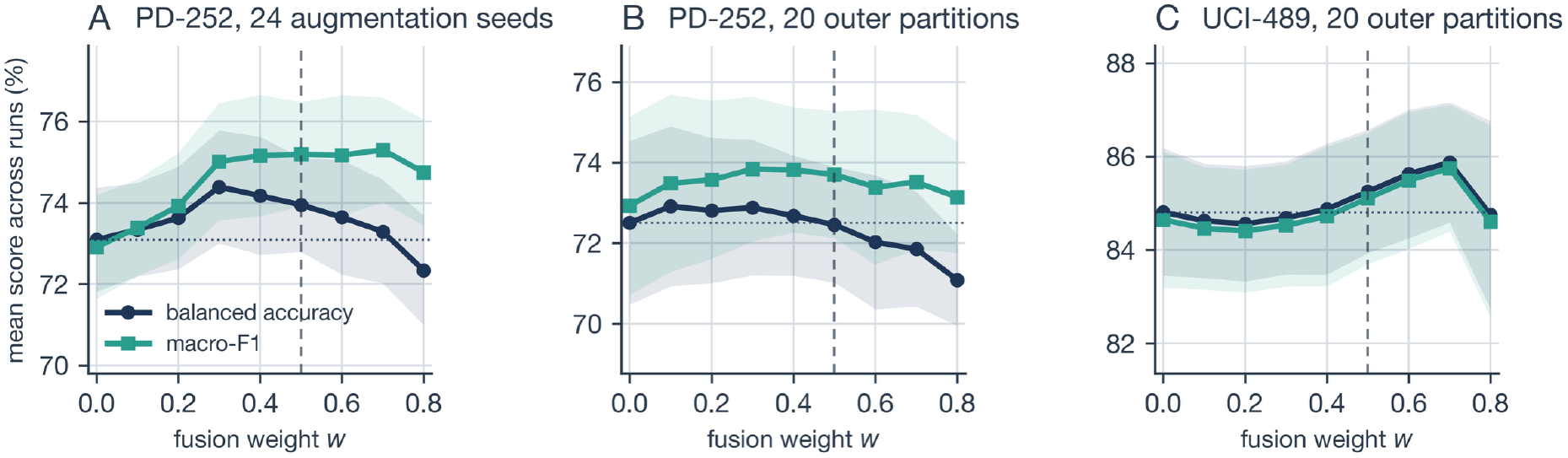
Sensitivity of the z-standardized full-space variant to the fusion weight *w* (Appendix A.3): mean balanced accuracy (navy circles) and macro-F1 (teal squares), in percent, across (**A**) the 24 augmentation seeds on PD-252, (**B**) the 20 outer partitions on PD-252, and (**C**) the 20 outer partitions on UCI-489. Shaded bands denote ± one standard deviation across runs, the dashed line marks the shared weight *w* = 0.5, and the dotted line marks the balanced accuracy of the matched Euclidean-kernel ablation (*w* = 0).

## Notes

### Competing Interest Statement

The authors have declared no competing interest.

